# Anatomical Determinants of Epilepsy Surgery Outcomes: A Systematic Review and Individual Patient Data Meta-Analysis

**DOI:** 10.1101/2025.05.13.25327534

**Authors:** Tamir Avigdor, Alyssa Ho, Matthew Moye, William Davalan, Erica Minato, Sana Hannan, Tamzin Holden, Tasha Bouchet, Yingqi Laetitia Wang, Kassem Jaber, Mays Khweileh, Samatha Kaplan, Vojtech Travnicek, David Carlson, Birgit Frauscher

## Abstract

**Importance:** To date, epilepsy surgery outcomes remain highly variable, with seizure freedom rates hovering around 50-70%, highlighting the need for a deeper understanding of the factors influencing surgical success.

**Objective:** To conduct an individual patient data meta-analysis of epilepsy surgery outcomes in drug-resistant epilepsy, leveraging granular, patient-level data to identify key clinical, demographic, and anatomical factors that influence surgical success.

**Data Sources:** MEDLINE (via Ovid), Embase, and Scopus were searched from inception to August 9, 2024.

**Study Selection:** Primary studies reporting patient-level surgical outcomes and clinical information in patients with drug-resistant epilepsy.

**Data Extraction and Synthesis:** Data were abstracted following the Preferred Reporting Items for Systematic Reviews and Meta-Analyses guidelines. Unique patient data from 385 studies were pooled, yielding 5,588 patients with outcomes, localization, demographics, pathology, and other findings. Surgical success rates were reported with 95% Wald confidence intervals.

**Main Outcome(s) and Measure(s):** Measured outcomes were surgical success rates (% Engel 1/ ILAE 1-2) based on key patient and disease-specific factors. Statistical associations were tested with chi-squared tests (p<0.05), effect sizes measured with Cramer’s V, and post-hoc comparisons adjusted using the false discovery rate.

**Results:** Surgical success rates (Engel I/ILAE 1-2) have remained stable over the past decades (r=0.25, p=0.13), while seizure freedom rates (Engel Ia/ILAE 1) have significantly improved (r=0.59, p<0.01). This occurred alongside a rise in surgical interventions, including more complex cases, as indicated by increased stereo-EEG use, and the adoption of minimally invasive techniques. Surgical success varied significantly by lobar anatomy (χ^2^=52, p<0.01), with the highest success rates in temporal (68.6% [67.0–70.1%]) and insular lobes (66.2% [55.4–77.0%]), although only temporal outcomes were statistically significant. Multilobar resections had lower success rates, with outcomes varying significantly by lobar combination (χ^2^=25, p=0.02). Variability in outcomes were also influenced by histopathology and MRI findings (χ^2^=121, p<0.001), and the type of surgical intervention (χ^2^=30.5, p<0.001).

**Conclusions and Relevance:** This meta-analysis combined patient-level data from multiple studies to understand how individual patient characteristics influence surgical outcomes.

Identifying these prognostic factors can guide more personalized patient selection and surgical planning, and ultimately improve rates of favorable outcomes in epilepsy surgery.

**Key Points:** *Question:* What are the main factors influencing surgical success in drug-resistant epilepsy patients?

*Findings:* A systematic review of 5,588 individual patient data from 385 primary research studies showed that the anatomical region, surgical technique, and histopathological diagnosis impact epilepsy surgery outcomes, with varying success rates based on these factors’ interaction.

*Meaning:* Presurgical evaluations and research into potential biomarkers and treatment options should consider these patient-specific factors instead of relying on generalized, population-level outcome statistics.

## Introduction

Epilepsy is one of the most prevalent and disabling neurological conditions, affecting individuals of all ages and impacting over 70 million individuals worldwide.^1^ Beyond its debilitating clinical effects, epilepsy carries with it significant comorbidities and substantial economic burden.^2^ In 2016, the World Health Organization’s Global Burden of Disease highlighted the significance of epilepsy, ranking it among the top neurological contributors to global disability-adjusted life-years.^2^

While antiseizure medication is the first line of treatment for epilepsy, nearly one-third of patients continue to experience seizures despite having been prescribed multiple and varied treatment regimens of antiseizure drugs in sufficient dosages.^3,4^ This condition, termed drug-resistant epilepsy^5^ poses a major challenge in epilepsy treatment, and is responsible for 80% of the total cost associated with the disorder.^6,7^ For these individuals, surgical intervention is a potentially life-changing option, offering the prospect of seizure freedom and improved quality of life.^8^

Epilepsy surgery can be divided into two main categories, resective and minimally invasive surgery, both aimed at removing the tissue indispensable for seizure generation (so-called “epileptogenic zone”),^9^ thereby aiming to render the patient seizure free.^8^ Resective surgery, the most invasive option, is typically used to remove larger volumes of diseased tissue, whereas ablation, encompassing laser interstitial thermal therapy (LITT), radiofrequency thermocoagulation, and focused ultrasound, offers a less invasive alternative.^10^

Despite advancements in epilepsy surgery,^11^ rates of post-surgical seizure freedom hover around 50-70% and are highly variable.^12^ Several factors contribute to this variability, such as limitations in electroencephalography (EEG)/intracranial-EEG (iEEG) monitoring for accurately capturing the epileptogenic zone,^13^ as well as differences in etiology, implicated brain regions,^14-16^ and demographic factors such as sex and age.^17^ Additionally, even in cases where epilepsy surgery may be successful, there is a severe underutilization of surgical interventions, with only 1% of eligible patients being referred for comprehensive epilepsy presurgical evaluations.^18,19^ This may stem from fear of complications and reservations about the benefits of surgery.^20^ These challenges highlight the importance of recognizing the personalized nature of epilepsy and the necessity of selecting tailored surgical interventions.

As such, there is a need to understand how different types of surgery, etiology, age, sex and specific brain regions interact and contribute to surgical success.^14-16^ Despite the known influences of these factors in affecting the likelihood of seizure freedom following surgery, their relative importance in determining surgical success remains to be determined. Many meta-analyses have examined seizure outcomes after epilepsy surgery, but they often focus on specific cohorts and rely on aggregate data, which may overlook key sources of heterogeneity. To address this, we conducted a comprehensive literature review and meta-analysis of individual patient data (IPD) to systematically quantify the rates of surgical success within a large international dataset of patients with drug-resistant epilepsy. As the gold standard for evaluating treatment effectiveness across diverse patient subgroups, IPD meta-analysis provides a more precise and nuanced assessment of surgical outcomes across various epilepsy and surgery types. This approach allowed us to identify key sources of heterogeneity between these rates, such as sex, age, and pathology, offering deeper insights into factors influencing surgical success. It also improves data handling, facilitates novel insights, and ensures greater transparency and reliability in findings. By integrating these findings, we aim to equip clinicians and patients with a more data-driven understanding of epilepsy surgery, thus optimizing patient outcomes and improving the accessibility and acceptance of surgical interventions in the management of drug-resistant epilepsy.

## Methods

### Search strategy and selection criteria

We conducted a systematic review to identify studies reporting patient-level data on epilepsy surgery outcomes in drug-resistant epilepsy for the purpose of an IPD meta-analysis. Our data collection and analyses followed a prespecified protocol (registered on Prospero on 17 April 2024: CRD42024530397).

The search strategy was developed in collaboration with a medical librarian (S.K.) who identified potential cohorts by searching OVID MEDLINE, Embase, and Scopus using a comprehensive search strategy (see Appendix 1). The search was conducted on August 9, 2024. The Covidence software was used for the review and management of studies. Title and abstract screening, as well as full-text reviews, were performed by two reviewers, with any conflicts resolved by a third reviewer. For the full list of inclusion and exclusion criteria, please see Supplementary Figure 1. Data extraction was performed by one reviewer and cross-checked by the first authors (T.A. or A.H.).

### Data extraction

For each patient, we extracted the following patient-level information: sex, age, pathology, type of epilepsy, affected hemisphere including brain region (if available), type of surgical intervention, region(s) resected, MRI information, and surgical outcome. Missing data was denoted as “unknown” and cross-checked for validation. To ensure consistency across studies with varied terminology, we standardized the format of each variable using established definitions and classifications defined by the ILAE whenever possible.^21-24^ Additionally, to mitigate duplicate entries from multiple papers sharing the same patient cohorts, we implemented a two-step screening process. First, an automated screening identified potential duplicates by matching key demographic and clinical variables, including sex, age, etiology, and center/country of origin. Suspected duplicates were then manually reviewed for confirmation before removal to ensure data integrity.

### Definition of surgical success

The definition of surgical success varies largely across studies. To standardize outcomes, we used two widely recognized measures. The Engel Outcome Scale^25^ classifies outcomes into four classes (Engel 1-4) based on the presence and improvement of disabling seizures, with Engel 1 indicating a favorable outcome and Engel 1a representing a completely seizure free outcome. The 2001 ILAE classification^26^ categorizes outcomes into six classes (ILAE 1-6) based on seizure frequency and semiology, with ILAE 1-2 indicating a favorable outcome, and ILAE 1 representing a seizure free outcome. For the main results of this meta-analysis, surgical success is defined as Engel 1 and ILAE 1-2, while supplementary analyses also report results associated with seizure-freedom using Engel 1a and ILAE 1.

### Parcellation

Cortical areas were grouped into five main lobes: temporal, parietal, insular, frontal, and occipital. Sublobar parcellation was performed using the Brainnetome Atlas,^27^ which provides a standardized framework for categorizing the epileptic focus and resected regions. All subsequent regional analyses were conducted based on these predefined groupings.

### Statistical Analysis

Surgical success rates were calculated as the percentage of patients in the positive group relative to those in the negative group, with 95% confidence intervals using Wald’s coefficient. Chi-squared tests were used to assess statistical associations between variables and surgical success in each sub-analysis, with significance set at p<0.05. Cramer’s V was used to measure the effect size. Post-hoc comparisons were adjusted for multiple corrections using the false discovery rate. We further explored data heterogeneity using a logistic regression model to estimate the predicted probability of surgical success based on all available parameters.

## Results

### Demographics and clinical characteristics

A total of 385 studies, providing patient-level data on 5,588 patients, met the selection criteria and were included in this meta-analysis (see inclusion flowchart, Supplementary Figure 1).^28-140^ Publication bias was assessed by a funnel plot for studies reporting patient-level data. Visual examination revealed mild asymmetry in the rates of surgical success (Supplementary Figure 2). A description of included papers is described in Supplementary Table 1.

Of the 5,588 patients, 2,220 were male, 2,129 were female, and sex was not specified for 1,239 patients. The most common epilepsy types in our cohort were temporal lobe epilepsy (n=3,252) and frontal lobe epilepsy (n=754). The median age at intervention was 24 years (0.5 to 77 years), with a median disease duration of 16.5 years (10 months to 60 years) between epilepsy diagnosis and surgical intervention. The median post-surgical follow-up interval was 24 months (12 to 480 months). The dataset represents patients from 36 countries, highlighting the global scope of this IPD meta-analysis (Supplementary Figure 3).

Overall, 3,894 patients (success rate 64.14% [95% CI 62.88 – 65.39%]) achieved a favorable surgical outcome (Engel 1 or ILAE 1-2) at the last follow-up. There were no significant differences between favorable and unfavorable surgical outcomes for sex, duration of disease, and post-surgical follow up time (p>0.05), while age at intervention showed a negligible effect (p=0.006, d=0.05). A summary of patient characteristics and corresponding rates of seizure freedom is presented in Figure 1.

**Figure 1.**
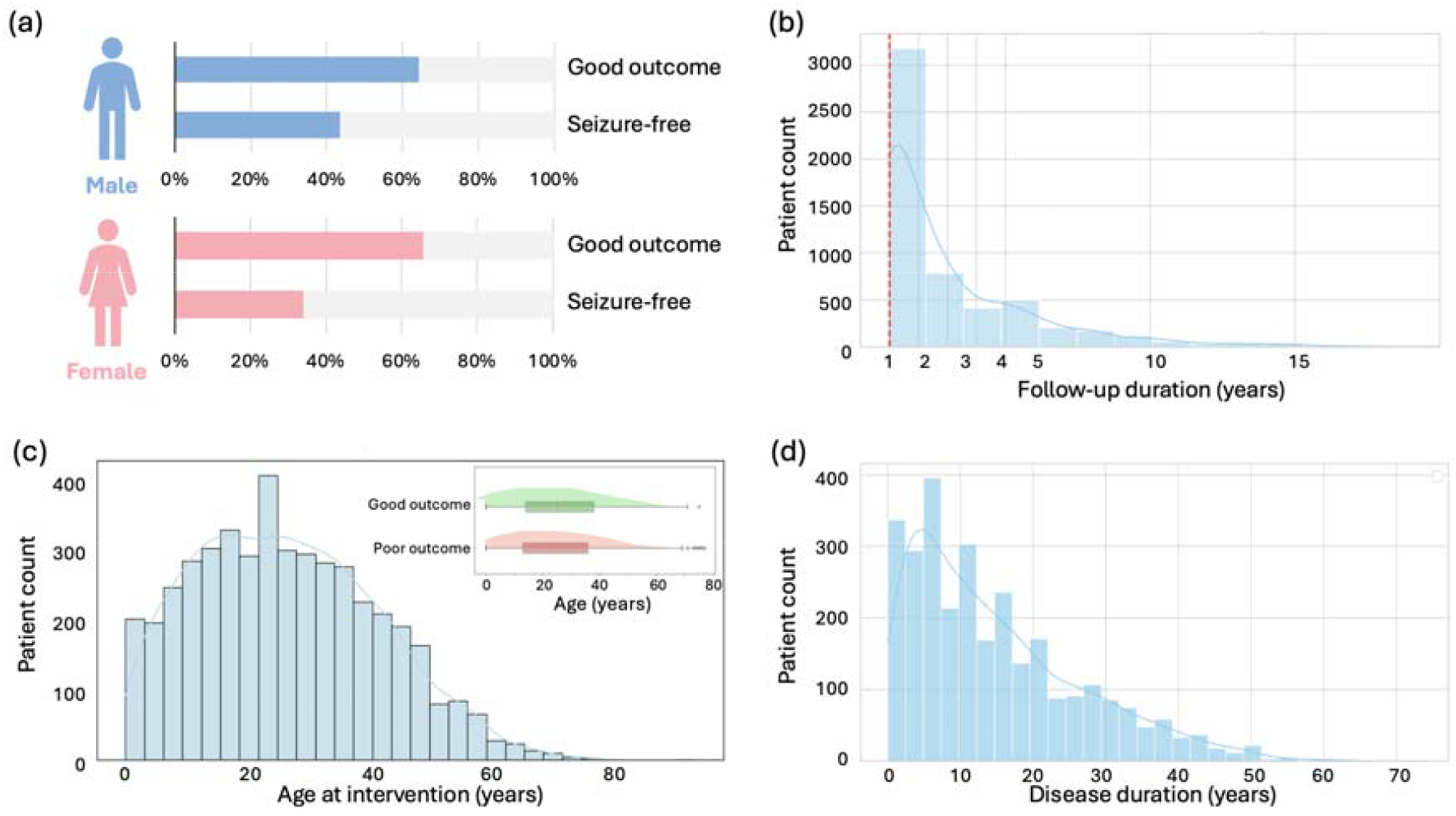
Summary of patient demographics. (a) Percentage of males and females with good outcomes and seizure-freedom. (b) Distribution of post-surgical follow-up duration in the cohort. (c) Distribution of patients and their age at time of surgical intervention. (d) Distribution of disease duration prior to surgical intervention.

### Type of surgical intervention

Surgical success rates (Engel I/ILAE 1-2) have remained stable over time (Figure 2a; r=0.25, p=0.13), while the rate of seizure freedom (Engel Ia/ILAE 1) has significantly improved (Supplementary Figure 4a). This trend persists despite the increasing adoption of minimally invasive surgeries and a growing number of patients undergoing surgical interventions (Figure 2b, Supplementary Figure 4b). There was a significant association between the type of surgical intervention and the rate of surgical success (χ^2^=30.5, p<0.001, V=0.07) (Figure 2c). Patients who underwent hemispherectomies (success rate 67.3% [95% CI 60.7 – 74.0%]) and resections (success rate 64.8% [95% CI 63.5 – 66.2%]) had a higher, albeit not statistically significant, success rate than those who received disconnective surgeries (success rate 60.9% [95% CI 40.9 – 80.8%]) or minimally invasive procedures such as LITT or radiofrequency thermocoagulation (success rate 60.5% [54.5 – 66.4%]). In contrast, corpus callosotomy (success rate 45.0% [95% CI 37.0 – 53.0%]), typically offered as a palliative surgery, had the lowest success rate, with comparisons with hemispherectomies, resections, and minimally invasive procedures reaching statistical significance (all p<0.001).

**Figure 2.**
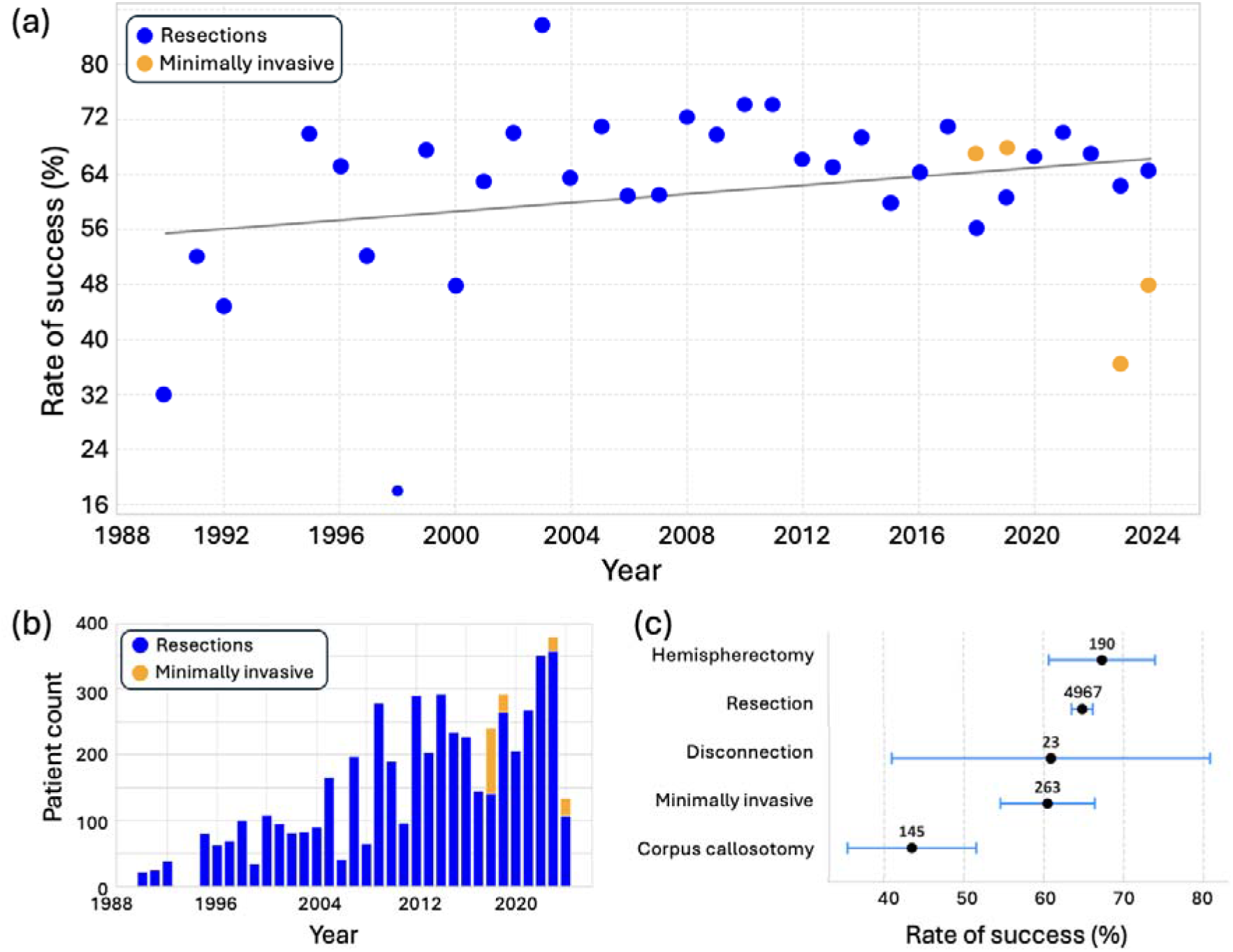
Type of surgical interventions. (a) Reported success rates of resections (blue), defined as Engel I or ILAE 1–2 outcomes, over time, alongside success rates for minimally invasive surgeries (yellow). Each dot represents the overall success rate calculated from all the patients pooled across studies conducted in a given year. Linear regression revealed no significant trend (r=0.25, p=0.13). (b) Number of patients undergoing resections and those undergoing minimally invasive surgery over time. Only years with ≥20 patients are shown. (c) Surgical success rates by intervention type. The highest success rate was observed in hemispherectomies (n=190, 67.4% [60.7–74.0%]), followed by resections (n=4,967, 64.8% [63.5–66.2%]), disconnections (n=23, 60.9% [40.9–80.8%]), and minimally invasive surgeries (n=263, 60.5% [54.5–66.4%]), with no significant differences among these groups (p>0.05). In contrast, success rates were significantly lower for corpus callosotomies (n=145, 43.4% [35.4–51.5%]) compared to hemispherectomies, resections, and minimally invasive surgeries (all p<0.01).

### Surgical success by brain region

We analyzed surgical success rates and rates of seizure freedom (Supplementary Figure 5) separately for patients who had procedures confined to a single lobe and those who underwent surgeries involving a combination of different lobes.

For unilobar surgeries, there was a significant association between the involved lobe and the rate of surgical success (χ^2^=52, p<0.001, V=0.12). Success rates were significantly higher in the temporal lobe (68.6% [67.0 – 70.1%]) compared to the occipital (55.8% [46.6 – 65.1%]), parietal (56.0% [49.0 – 63.0%]), and frontal lobe (57.3% [53.8-60.8%]) (all p<0.05) (Figure 3a). The insula demonstrated the second highest success rate at 66.2% [55.4–77.0%], although this was not statistically significant. Within the temporal lobe, we further conducted a sublobar analysis (Figure 3b), revealing no significant difference between surgeries of the mesial temporal lobe (63.1% [59.0 – 67.1%]) and those of the lateral and basal temporal lobe (71.0% [62.1 – 80.0%]).

**Figure 3.**
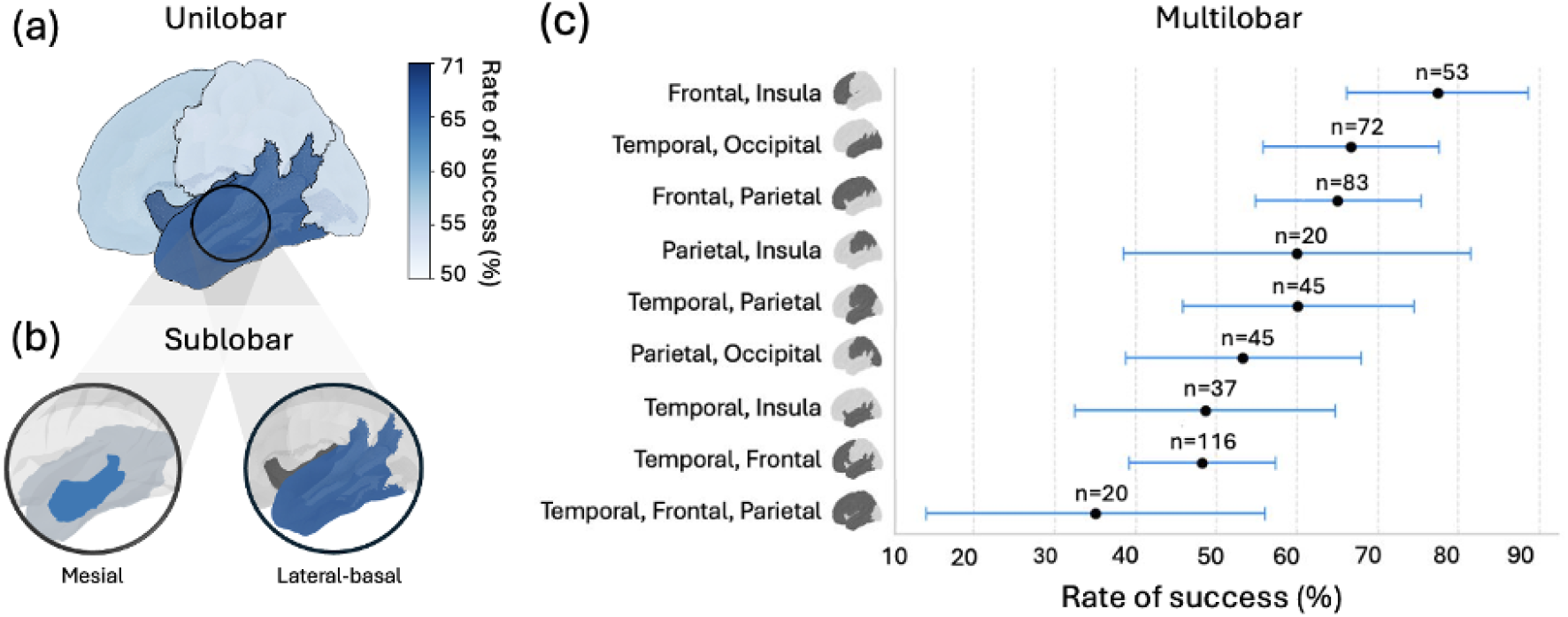
Surgical success rates across unilobar (n=4384), sublobar (n=655) and multilobar (n=491) surgeries. (a) For unilobar surgeries, surgical success significantly varied by lobe (χ^2^=52, p<0.001, V=0.12). Success rates were as follows: temporal lobe (n=3,252, 68.6% [67.0–70.2%]), insula (n=74, 66.2% [55.4–77.0%]), frontal lobe (n=754, 57.3% [53.8– 60.8%]), parietal lobe (n=193, 56.0% [49.0–63.0%]), and occipital lobe (n=111, 55.9% [46.6–65.1%]). Only the differences between the temporal and frontal lobe, temporal and parietal lobe and temporal and occipital lobe were significant (p<0.01). (b) Within the temporal lobe, sublobar analysis showed that mesial temporal surgeries (n=555) had a success rate of 63.1% (59.0–67.1%), whereas procedures targeting the lateral and basal temporal regions (n=100) had a success rate of 71.0% (62.1–79.9%). This difference was not significantly different. (c) Multilobar surgeries involved combinations of the frontal, temporal, insular, and parietal lobes. There was a significant association between surgical region and success rate (χ^2^=25, p=0.02, V=0.22). Success rates for different multilobar combinations were: frontal-insula (77.4% [66.1–88.6%]), temporal-occipital (66.7% [55.8– 77.6%]), frontal-parietal (65.1% [54.8–75.3%]), temporal-parietal (60.0% [45.7–74.3%]), parietal-insula (60.0% [38.5–81.5%]), parietal-occipital (53.3% [38.8–67.9%]), temporal-insula (48.6% [32.5–64.8%]), temporal-frontal (48.3% [39.2–57.4%]), and temporal-frontal-parietal (35.0% [14.1–55.9%]). Significant differences were observed between the following pairs: temporal-frontal vs. temporal-occipital (p=0.02, V=0.17), temporal-frontal vs. frontal-parietal (p=0.03, V=0.16), temporal-occipital vs. temporal-frontal-parietal (p=0.02, V=0.24), and frontal-parietal vs. temporal-frontal-parietal (p=0.03, V=0.22).

For multilobar surgeries (Figure 3c), the procedures performed included combinations involving the frontal, temporal, insular, and parietal lobes. Overall, there was a significant association between the surgical region and success rate (χ^2^=25, p=0.02, V=0.22). Surgeries involving the frontal and insular lobes had the highest success rate (77.4% [66.1 – 88.6%]). In contrast, procedures affecting the temporal, frontal and parietal lobes had the lowest success rate (35.0% [14.1 – 55.9%]). This difference was statistically significant (p=0.03, V=0.22).

### Direct surgery vs. iEEG-informed surgery

The use of iEEG, particularly stereo-EEG, has expanded globally and gained popularity over the past decade,^141^ becoming an integral tool for improving the localization of the epileptic focus, particularly for more complex cases (Figure 4a and 4b). Surgical success rates for patients who had SEEG did not change significantly over time (n=1330, p=0.27). Among unilobar cases (Figure 4c), surgical success varied significantly by lobe (χ^2^=6.2, p=0.004, V=0.11). These trends were consistent with those observed in the overall cohort.

**Figure 4.**
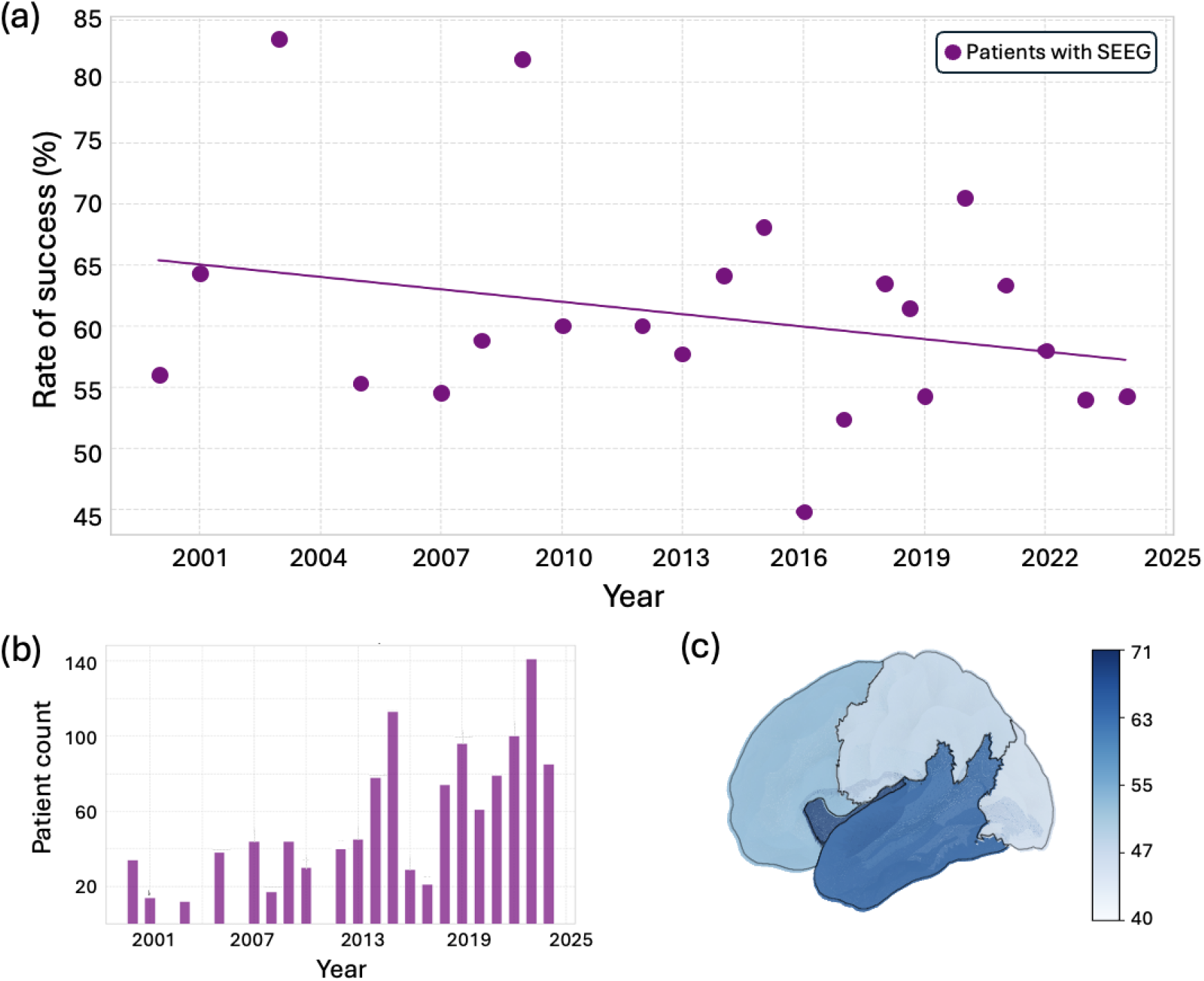
Utilization of stereo-electroencephalography (SEEG) in presurgical evaluation and its impact on surgical outcomes. (a) Surgical success rates among patients who underwent SEEG remained relatively stable over time, with no significant trend observed (p = 0.27). (b) The number of patients receiving SEEG as part of their presurgical evaluation increased over the years. (c) In unilobar cases, surgical success rates varied significantly by lobe (p = 0.18, V = 0.08), with the highest success in the insula (69.2% [44.1-94.3%]) and temporal lobe (64.2% [60.4-68.1%]), followed by the frontal (54.9% [48.9-60.9%]), parietal (49.1% [35.6-62.5%]), and occipital lobes (47.6% [26.3-69.0%]). The differences in surgical success rates between the temporal and frontal lobes (p = 0.004, V = 0.11) and between the temporal and parietal lobes (p = 0.02, V = 0.11) were statistically significant.

### Surgical success by pathology

Subgroup analyses were performed based on pathology, with malformations of cortical development (n=1,340), hippocampal sclerosis (n=950), tumors (n=588), glial scar (n=397), and vascular malformation (n=188) identified as the most common diagnoses in our cohort (Figure 5a). There was a significant association between the type of pathology and the rate of surgical success (χ^2^=121, p<0.001, V=0.16). Surgical success was highest in patients with tumors (81.6% [74.9 – 81.6%]) and lowest in those with glial scars (56.3% [46.5 – 56.3%]). This difference was statistically significant (p<0.01, V=0.28).

**Figure 5.**
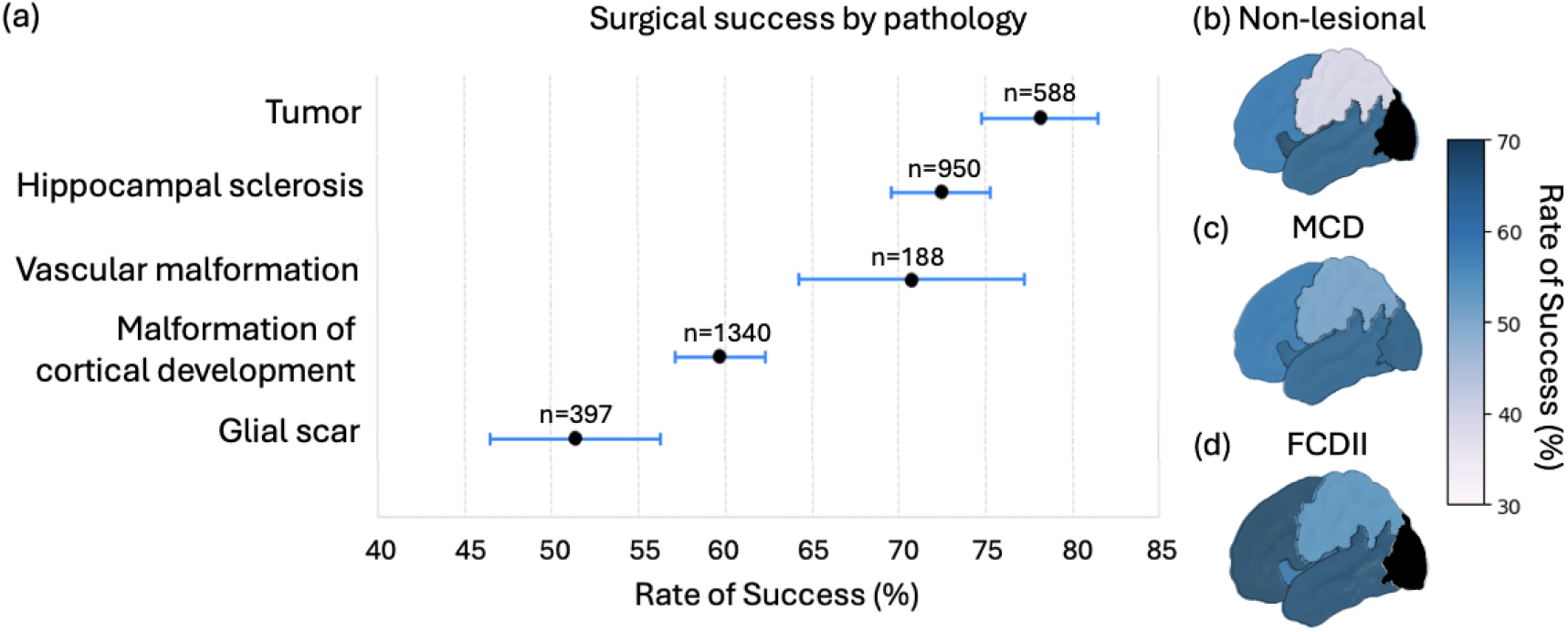
Surgical success rates by pathology. Pathology was determined by MRI, histology, or both. (a) Success rates for the top five pathologies were: tumors (78.2% [74.9–81.6%]), hippocampal sclerosis (72.5% [69.7–75.4%]), vascular malformation (70.7% [64.2–77.2%]), malformation of cortical development (59.7% [57.1–62.3%]), and glial scars (51.4% [46.5– 56.3%]). All paired comparisons were statistically significant (p<0.01), except for hippocampal sclerosis vs. vascular malformation (p=0.68, V=0.01). (b) For MRI negative epilepsy, success rate by lobe was: insula (n=23, 65.2% [45.8–84.7%]), temporal lobe (n=341, 58.1% [52.8–63.3%]), frontal lobe (n=139, 57.6% [49.3–65.8%]), and parietal lobe (38.7% [21.6–55.9%]). Only the difference between the temporal and parietal lobes was statistically significant (p=0.02, V=0.13). (c) In malformation of cortical development (MCD), success rates by lobe were: occipital (n=36, 66.7% [51.3–82.1%]), insula (n=40, 65.0% [50.2–79.8%]), temporal (n=497, 65.4% [61.2–69.6%]), frontal (n=385, 60.3% [55.4– 65.1%]), and parietal (n=86, 52.3% [41.8–62.9%]). The only statistically significant difference was between the temporal and parietal lobes (p=0.03, V=0.09). (d) For focal cortical dysplasia (FCD) type II, there was no significant association between lobe and success rate (p=0.18, V=0.08). Success rates by lobe was: temporal (n=162, 69.1% [62.0– 76.2%]), frontal (n=171, 69.0% [62.1–75.9%]), insula (n=22, 59.1% [38.5–79.6%]), and parietal (n=45, 55.6% [41.0–70.1%]). Black regions indicate insufficient sample size, defined as n<20.

For “MRI-negative” epilepsy (Figure 5b), surgical success was significantly associated with the brain region (χ^2^=26, p=0.02, V=0.23), with the temporal lobe (58.1% [52.8-63.3%]) having a significantly higher success rate compared to the parietal lobe (38.7% [21.6 – 55.9%]) (p=0.02, V=0.13). Similarly, in patients with malformations of cortical development, surgical success was significantly associated with the brain region (χ^2^=26, p=0.03, V=0.22) (Figure 5c). Resections targeting malformations in the temporal lobe (65.4% [61.2-69.6%]) had a significantly higher success rate compared to those in the parietal lobe (52.3% [41.8 – 62.9%]). There was no significant association between brain regions and surgical success in cases of focal cortical dysplasia (FCD) type II (χ^2^=6.2, p=0.18, V=0.08) (Figure 5d). Results for seizure freedom by pathology are presented in Supplementary Figure 6.

For the mesial temporal lobe, surgical success rates varied by pathology type (p<0.006, V=0.21). Rates were highest for hippocampal sclerosis (n=172; 70%) and lowest for glial scars (n=22; 55%) (Supplementary Figure 7). These differences were not significant.

## Discussion

This study presents a comprehensive IPD meta-analysis of surgical outcomes in drug-resistant epilepsy, incorporating data from 385 studies and 5,588 patients. Our findings highlight that surgical success varies based on the affected brain region, type of surgical intervention, and underlying pathology, whereas there was no significant association between surgical outcomes and patient sex, disease duration, and post-surgical follow up duration.

While there have been numerous meta-analyses exploring seizure outcomes following epilepsy surgery, they often focus on specific epilepsy types or patient cohorts,^142-144^ and primarily rely on aggregate data.^145,146^ However, IPD meta-analyses have been shown to produce different results in 20% of cases and are 15% more likely to identify statistically significant differences that aggregated analyses may overlook.^147^ This is particularly relevant for epilepsy surgery outcomes, given its diverse etiology and the persistent variability in surgical outcomes. Indeed, despite technological and diagnostic advancements over the years, seizure freedom rates remain between 50–70%, with long-term prognosis often being even less favorable.^12,145^

Our analysis highlights the notably high success rates of insular lobe surgeries, particularly when guided by SEEG. This challenges conventional expectations and suggests that the insula may be a more favorable surgical target than previously recognized.^148,149^ Insular lobe epilepsy has long been considered difficult to diagnose due to its reputation as a “great mimicker” of other focal epilepsies and its poor visibility in non-invasive investigations.^149,150^ Indeed, it was the least frequently performed unilobar surgery in our cohort. However, with increased SEEG sampling of the insula, more patients are being accurately diagnosed and successfully treated with surgery, leading to improved outcomes.^151,152^ Similarly, temporal lobe surgeries continued to demonstrate high rates of favorable outcomes, reinforcing their well-established role as an effective surgical target.^142,151^ In contrast, surgeries involving the posterior cortex, including the occipital and parietal lobes, were associated with the lowest success rates. This is likely due to the functional significance of both the occipital and parietal lobes.^40,135,153^ The occipital lobe is crucial for visual processing,^138,139^ and the parietal lobe is involved in spatial and sensory integration,^40,135,140^ making surgical resection more complex and less likely to be fully successful. Both lobes present challenges in achieving complete resection while preserving function. Given these findings, increased awareness and refined surgical approaches for insular and posterior cortex epilepsies are warranted. Expanding the use of SEEG in evaluating complex cases may further improve patient selection and surgical outcomes, particularly for insular epilepsy, where growing evidence supports its viability as a surgical target.

Our results further show that patients with malformations of cortical development, particularly those with FCD type II, achieve more favorable surgical outcomes. This aligns with recent findings from the MELD consortium,^16^ reinforcing FCD type II as a strong prognostic marker for surgical success. Given these promising outcomes, surgery should be more readily considered for this patient group, as it offers a high likelihood of seizure freedom and represents a highly effective treatment option. These findings are also consistent with recommendations from the Surgical Therapies Commission of the ILAE, which, through a Delphi consensus process involving 61 experts, advocated for broader surgical referral criteria.^154^ Among their recommendations, they emphasized considering surgery even for non–drug-resistant patients with brain lesions in non-eloquent cortex,^154^ highlighting the strong prognostic value of pathological indicators identified through MRI. This expert consensus supports the growing body of evidence promoting earlier and more widespread consideration of epilepsy surgery to optimize patient outcomes.

### Strengths and limitations

Our meta-analysis has several key strengths, including its rigorous methodology and the use of individual-level patient data from a large international cohort of patients with drug-resistant epilepsy. This approach allowed for a more precise and nuanced evaluation of surgical success across diverse epilepsy types and procedures. Additionally, we meticulously screened the data and conducted thorough tests for publication bias, ensuring the robustness and reliability of our findings. Despite these strengths, several challenges arose in pooling data due to variability in the level of detail reported across studies and the lack of standardization in published data. To mitigate this, we applied standardized terminology defined by the ILAE whenever possible.^21-24^ However, the inconsistencies between studies highlight the need for greater uniformity in reporting. Future large-scale, prospective, multicenter studies with standardized methodologies are essential to refine our understanding of surgical outcomes and enhance the ability to model disease trajectories more accurately.

## Conclusions

In summary, the findings of this IPD meta-analysis suggest key factors associated with surgical success, including age at intervention, the type of surgical approach. Moreover, the observed variability in outcomes by anatomical region may inspire the broader consideration of surgical interventions for epilepsy types traditionally associated with poor prognoses, such as insular lobe epilepsy. These insights can aid clinicians and patients in presurgical decision-making and risk counseling.

## Data Availability

All data produced are available online at surveyed papers

## Acknowledgments

We would like to thank Arpana Wadhwani and Marah Abdelkader for their valuable assistance with the title and abstract screening of manuscripts.

## References

1 Ngugi, A. K., Bottomley, C., Kleinschmidt, I., Sander, J. W. & Newton, C. R. Estimation of the burden of active and life-time epilepsy: a meta-analytic approach. Epilepsia 51, 883–890 (2010). 10.1111/j.1528-1167.2009.02481.x

2 Collaborators, G. B. D. E. Global, regional, and national burden of epilepsy, 1990-2016: a systematic analysis for the Global Burden of Disease Study 2016. Lancet Neurol 18, 357–375 (2019). 10.1016/S1474-4422(18)30454-X

3 Kalilani, L., Sun, X., Pelgrims, B., Noack-Rink, M. & Villanueva, V. The epidemiology of drug-resistant epilepsy: A systematic review and meta-analysis. Epilepsia 59, 2179–2193 (2018). 10.1111/epi.14596

4 Chen, Z., Brodie, M. J., Liew, D. & Kwan, P. Treatment Outcomes in Patients With Newly Diagnosed Epilepsy Treated With Established and New Antiepileptic Drugs: A 30-Year Longitudinal Cohort Study. JAMA Neurol 75, 279–286 (2018). 10.1001/jamaneurol.2017.3949

5 Kwan, P. et al. Definition of drug resistant epilepsy: consensus proposal by the ad hoc Task Force of the ILAE Commission on Therapeutic Strategies. Epilepsia 51, 1069–1077 (2010). 10.1111/j.1528-1167.2009.02397.x

6 Engel, J., Jr. et al. Early surgical therapy for drug-resistant temporal lobe epilepsy: a randomized trial. JAMA 307, 922–930 (2012). 10.1001/jama.2012.220

7 Begley, C. E. et al. The cost of epilepsy in the United States: an estimate from population-based clinical and survey data. Epilepsia 41, 342–351 (2000). 10.1111/j.1528-1157.2000.tb00166.x

8 Ryvlin, P., Cross, J. H. & Rheims, S. Epilepsy surgery in children and adults. Lancet Neurol 13, 1114–1126 (2014). 10.1016/S1474-4422(14)70156-5

9 Rosenow, F. & Luders, H. Presurgical evaluation of epilepsy. Brain 124, 1683–1700 (2001). 10.1093/brain/124.9.1683

10 Hoppe, C. et al. Laser interstitial thermotherapy (LiTT) in epilepsy surgery. Seizure 48, 45–52 (2017). 10.1016/j.seizure.2017.04.002

11 Medina-Pizarro, M., Spencer, D. D. & Damisah, E. C. Recent advances in epilepsy surgery. Curr Opin Neurol 36, 95–101 (2023). 10.1097/WCO.0000000000001134

12 de Tisi, J. et al. The long-term outcome of adult epilepsy surgery, patterns of seizure remission, and relapse: a cohort study. Lancet 378, 1388–1395 (2011). 10.1016/S0140-6736(11)60890-8

13 Minotti, L., Montavont, A., Scholly, J., Tyvaert, L. & Taussig, D. Indications and limits of stereoelectroencephalography (SEEG). Neurophysiol Clin 48, 15–24 (2018). 10.1016/j.neucli.2017.11.006

14 Englot, D. J. et al. Seizure types and frequency in patients who “fail” temporal lobectomy for intractable epilepsy. Neurosurgery 73, 838-844; quiz 844 (2013). 10.1227/NEU.0000000000000120

15 Englot, D. J. et al. Factors associated with failed focal neocortical epilepsy surgery. Neurosurgery 75, 648-645;discussion 655; quiz 656 (2014). 10.1227/NEU.0000000000000530

16 Wagstyl, K. et al. Atlas of lesion locations and postsurgical seizure freedom in focal cortical dysplasia: A MELD study. Epilepsia 63, 61–74 (2022). 10.1111/epi.17130

17 Bjellvi, J., Edelvik Tranberg, A., Rydenhag, B. & Malmgren, K. Risk Factors for Seizure Worsening After Epilepsy Surgery in Children and Adults: A Population-Based Register Study. Neurosurgery 87, 704–711 (2020). 10.1093/neuros/nyz488

18 Okubo, Y., Fallah, A., Hayakawa, I., Handa, A. & Nariai, H. Trends in hospitalization and readmission for pediatric epilepsy and underutilization of epilepsy surgery in the United States. Seizure 80, 263–269 (2020). 10.1016/j.seizure.2020.05.013

19 Haneef, Z., Stern, J., Dewar, S. & Engel, J., Jr. Referral pattern for epilepsy surgery after evidence-based recommendations: a retrospective study. Neurology 75, 699–704 (2010). 10.1212/WNL.0b013e3181eee457

20 Culler, G. W. t. & Jobst, B. C. Surgical Treatments for Epilepsy. Continuum (Minneap Minn) 28, 536–558 (2022). 10.1212/CON.0000000000001106

21 Blumcke, I. et al. Histopathological Findings in Brain Tissue Obtained during Epilepsy Surgery. N Engl J Med 377, 1648–1656 (2017). 10.1056/NEJMoa1703784

22 Blumcke, I. et al. The clinicopathologic spectrum of focal cortical dysplasias: a consensus classification proposed by an ad hoc Task Force of the ILAE Diagnostic Methods Commission. Epilepsia 52, 158–174 (2011). 10.1111/j.1528-1167.2010.02777.x

23 Fisher, R. S. et al. Operational classification of seizure types by the International League Against Epilepsy: Position Paper of the ILAE Commission for Classification and Terminology. Epilepsia 58, 522–530 (2017). 10.1111/epi.13670

24 Scheffer, I. E. et al. ILAE classification of the epilepsies: Position paper of the ILAE Commission for Classification and Terminology. Epilepsia 58, 512–521 (2017). 10.1111/epi.13709

25 Engel, J., Jr. Update on surgical treatment of the epilepsies. Summary of the Second International Palm Desert Conference on the Surgical Treatment of the Epilepsies (1992). Neurology 43, 1612–1617 (1993). 10.1212/wnl.43.8.1612

26 Wieser, H. G. et al. ILAE Commission Report. Proposal for a new classification of outcome with respect to epileptic seizures following epilepsy surgery. Epilepsia 42, 282–286 (2001).

27 Fan, L. et al. The Human Brainnetome Atlas: A New Brain Atlas Based on Connectional Architecture. Cereb Cortex 26, 3508–3526 (2016). 10.1093/cercor/bhw157

28 Abdallah, C. et al. Clinical Yield of Electromagnetic Source Imaging and Hemodynamic Responses in Epilepsy: Validation With Intracerebral Data. Neurology 98, e2499–e2511 (2022). 10.1212/WNL.0000000000200337

29 Abel, T. J. et al. Frameless robot-assisted stereoelectroencephalography in children: technical aspects and comparison with Talairach frame technique. J Neurosurg Pediatr 22, 37–46 (2018). 10.3171/2018.1.PEDS17435

30 Aghakhani, Y. et al. The role of periventricular nodular heterotopia in epileptogenesis. Brain 128, 641–651 (2005). 10.1093/brain/awh388

31 Akeret, K. et al. Ultrasonographic features of focal cortical dysplasia and their relevance for epilepsy surgery. Neurosurg Focus 45, E5 (2018). 10.3171/2018.6.FOCUS18221

32 Alsaadi, T. M. et al. Potentially misleading extratemporal lobe lesions in patients with temporal lobe epilepsy. J Neurol Neurosurg Psychiatry 74, 566–569 (2003). 10.1136/jnnp.74.5.566

33 Alshafai, L. et al. Clinical, EEG, MRI, MEG, and surgical outcomes of pediatric epilepsy with astrocytic inclusions versus focal cortical dysplasia. Epilepsia 55, 1568–1575 (2014). 10.1111/epi.12756

34 Arevalo-Astrada, M. A. et al. Epilepsy surgery in stroke-related epilepsy. Seizure 88, 116–124 (2021). 10.1016/j.seizure.2021.04.002

35 Aung, T. et al. The feasibility and value of extraoperative and adjuvant intraoperative stereoelectroencephalography in rolandic and perirolandic epilepsies. J Neurosurg Pediatr 27, 36–46 (2021). 10.3171/2020.6.PEDS2099

36 Aznarez, P. B., Cabeza, M. P., Quintana, A. S. A., Lara-Almunia, M. & Sanchez, J. A. Evolution of patients with surgically treated drug-resistant occipital lobe epilepsy. Surg Neurol Int 11, 222 (2020). 10.25259/SNI_251_2020

37 Bakhtiar, Y. et al. Posttraumatic epilepsy: A single institution case series in Indonesia. Surg Neurol Int 13, 298 (2022). 10.25259/SNI_142_2022

38 Bansal, L. et al. PET hypermetabolism in medically resistant childhood epilepsy: Incidence, associations, and surgical outcome. Epilepsia 57, 436–444 (2016). 10.1111/epi.13311

39 Barba, C. et al. Epilepsy surgery in Neurofibromatosis Type 1. Epilepsy Res 105, 384–395 (2013). 10.1016/j.eplepsyres.2013.02.021

40 Bartolomei, F. et al. Neural networks underlying parietal lobe seizures: a quantified study from intracerebral recordings. Epilepsy Res 93, 164–176 (2011). 10.1016/j.eplepsyres.2010.12.005

41 Bauer, R. et al. Outcome of adult patients with temporal lobe tumours and medically refractory focal epilepsy. Acta Neurochir (Wien) 149, 1211-1216; discussion 1216-1217 (2007). 10.1007/s00701-007-1366-z

42 Benedetti-Isaac, J. C. et al. Long-term surgical outcomes in patients with drug-resistant temporal lobe epilepsy and no histological abnormalities. Neurologia 28, 543–549 (2013). 10.1016/j.nrl.2013.01.011

43 Benifla, M. et al. Temporal lobe surgery for intractable epilepsy in children: What to do with the hippocampus? Seizure 52, 81–88 (2017). 10.1016/j.seizure.2017.09.020

44 Benifla, M. et al. Neurosurgical management of intractable rolandic epilepsy in children: role of resection in eloquent cortex. Clinical article. J Neurosurg Pediatr 4, 199–216 (2009). 10.3171/2009.3.PEDS08459

45 Blauwblomme, T. et al. Prognostic value of insular lobe involvement in temporal lobe epilepsy: a stereoelectroencephalographic study. Epilepsia 54, 1658–1667 (2013). 10.1111/epi.12260

46 Boling, W. Minimal access keyhole surgery for mesial temporal lobe epilepsy. J Clin Neurosci 17, 1180–1184 (2010). 10.1016/j.jocn.2010.01.030

47 Boling, W., Aghakhani, Y., Andermann, F., Sziklas, V. & Olivier, A. Surgical treatment of independent bitemporal lobe epilepsy defined by invasive recordings. J Neurol Neurosurg Psychiatry 80, 533–538 (2009). 10.1136/jnnp.2008.155291

48 Bourgeois, M. et al. Surgical treatment of epilepsy in Sturge-Weber syndrome in children. J Neurosurg 106, 20–28 (2007). 10.3171/ped.2007.106.1.20

49 Buchfelder, M., Fahlbusch, R., Ganslandt, O., Stefan, H. & Nimsky, C. Use of intraoperative magnetic resonance imaging in tailored temporal lobe surgeries for epilepsy. Epilepsia 43, 864–873 (2002). 10.1046/j.1528-1157.2002.46201.x

50 Buraniqi, E. et al. Temporal Encephalocele: A Treatable Etiology of Drug-Resistant Pediatric Temporal Lobe Epilepsy. Pediatr Neurol 142, 32–38 (2023). 10.1016/j.pediatrneurol.2022.12.015

51 Burneo, J. G. et al. Temporal lobectomy in congenital porencephaly associated with hippocampal sclerosis. Arch Neurol 60, 830–834 (2003). 10.1001/archneur.60.6.830

52 Busch, R. M. et al. Poor presurgical performance on both verbal and visual memory measures is associated with low risk for memory decline following left temporal lobectomy for intractable epilepsy. Epileptic Disord 10, 199–205 (2008). 10.1684/epd.2008.0199

53 Caicoya, A. G., Macarron, J., Albisua, J. & Serratosa, J. M. Tailored resections in occipital lobe epilepsy surgery guided by monitoring with subdural electrodes: characteristics and outcome. Epilepsy Res 77, 1–10 (2007). 10.1016/j.eplepsyres.2007.07.004

54 Cajigas, I. et al. Magnetic Resonance-Guided Laser Interstitial Thermal Therapy for Mesial Temporal Epilepsy: A Case Series Analysis of Outcomes and Complications at 2-Year Follow-Up. World Neurosurg 126, e1121–e1129 (2019). 10.1016/j.wneu.2019.03.057

55 Cao, Q. et al. The localization and lateralization of fear aura and its surgical prognostic value in patients with focal epilepsy. Ann Clin Transl Neurol 9, 1116–1124 (2022). 10.1002/acn3.51607

56 Carreno, M. et al. Epilepsy surgery in drug resistant temporal lobe epilepsy associated with neuronal antibodies. Epilepsy Res 129, 101–105 (2017). 10.1016/j.eplepsyres.2016.12.010

57 Casadei, C. H. et al. All-cause mortality and SUDEP in a surgical epilepsy population. Epilepsy Behav 108, 107093 (2020). 10.1016/j.yebeh.2020.107093

58 Casciato, S. et al. Epilepsy surgery in adult-onset Rasmussen’s encephalitis: case series and review of the literature. Neurosurg Rev 38, 463-470; discussion 470-461 (2015). 10.1007/s10143-015-0623-5

59 Catarino, C. B. et al. Genomic microdeletions associated with epilepsy: not a contraindication to resective surgery. Epilepsia 52, 1388–1392 (2011). 10.1111/j.1528-1167.2011.03087.x

60 Centeno, M. et al. Combined electroencephalography-functional magnetic resonance imaging and electrical source imaging improves localization of pediatric focal epilepsy. Ann Neurol 82, 278–287 (2017). 10.1002/ana.25003

61 Chandra, P. S. et al. Hemispherotomy for intractable epilepsy. Neurol India 56, 127–132 (2008).

62 Chang, E. F., Nagarajan, S. S., Mantle, M., Barbaro, N. M. & Kirsch, H. E. Magnetic source imaging for the surgical evaluation of electroencephalography-confirmed secondary bilateral synchrony in intractable epilepsy. J Neurosurg 111, 1248–1256 (2009). 10.3171/2009.6.JNS081376

63 Chassoux, F. Stereo-EEG: the Sainte-Anne experience in focal cortical dysplasias. Epileptic Disord 5 Suppl 2, S95–103 (2003).

64 Chen, H. H. et al. Cognitive and epilepsy outcomes after epilepsy surgery caused by focal cortical dysplasia in children: early intervention maybe better. Childs Nerv Syst 30, 1885–1895 (2014). 10.1007/s00381-014-2463-y

65 Chen, P. C., Baumgartner, J., Seo, J. H., Korostenskaja, M. & Lee, K. H. Bilateral intracranial EEG with corpus callosotomy may uncover seizure focus in nonlocalizing focal epilepsy. Seizure 24, 63–69 (2015). 10.1016/j.seizure.2014.08.011

66 Chen, X. et al. Predictive value of electrocorticography in epilepsy patients with unilateral hippocampal sclerosis undergoing selective amygdalohippocampectomy. Neurosurg Rev 29, 108–113 (2006). 10.1007/s10143-005-0002-8

67 Choi, J. Y. et al. Extratemporal hypometabolism on FDG PET in temporal lobe epilepsy as a predictor of seizure outcome after temporal lobectomy. Eur J Nucl Med Mol Imaging 30, 581–587 (2003). 10.1007/s00259-002-1079-8

68 Chugani, H. T. et al. “Subtotal” hemispherectomy in children with intractable focal epilepsy. Epilepsia 55, 1926–1933 (2014). 10.1111/epi.12845

69 Cimbalnik, J. et al. Multi-feature localization of epileptic foci from interictal, intracranial EEG. Clin Neurophysiol 130, 1945–1953 (2019). 10.1016/j.clinph.2019.07.024

70 Citherlet, D. et al. Sensory profile alterations in patients with insular epilepsy surgery: Preliminary findings. Epilepsy Behav 115, 107499 (2021). 10.1016/j.yebeh.2020.107499

71 Cohen-Gadol, A. A. et al. Nonlesional central lobule seizures: use of awake cortical mapping and subdural grid monitoring for resection of seizure focus. J Neurosurg 98, 1255–1262 (2003). 10.3171/jns.2003.98.6.1255

72 Cossu, M. et al. Stereoelectroencephalography in the presurgical evaluation of children with drug-resistant focal epilepsy. J Neurosurg 103, 333–343 (2005). 10.3171/ped.2005.103.4.0333

73 Cossu, M. et al. Stereo-EEG-guided radio-frequency thermocoagulations of epileptogenic grey-matter nodular heterotopy. J Neurol Neurosurg Psychiatry 85, 611–617 (2014). 10.1136/jnnp-2013-305514

74 Costa, E. et al. The Anterior Trans-Superior Temporal Gyrus Approach for Selective Amygdalohippocampectomy. World Neurosurg 159, e244–e251 (2022). 10.1016/j.wneu.2021.12.034

75 D’Agostino, M. D. et al. Posterior quadrantic dysplasia or hemi-hemimegalencephaly: a characteristic brain malformation. Neurology 62, 2214–2220 (2004). 10.1212/01.wnl.0000130459.91445.91

76 D’Alessio, L. et al. Psychiatric outcome of epilepsy surgery in patients with psychosis and temporal lobe drug-resistant epilepsy: a prospective case series. Epilepsy Behav 37, 165–170 (2014). 10.1016/j.yebeh.2014.06.002

77 Daniel, R. T., Meagher-Villemure, K., Farmer, J. P., Andermann, F. & Villemure, J. G. Posterior quadrantic epilepsy surgery: technical variants, surgical anatomy, and case series. Epilepsia 48, 1429–1437 (2007). 10.1111/j.1528-1167.2007.01095.x

78 Danielsson, S., Rydenhag, B., Uvebrant, P., Nordborg, C. & Olsson, I. Temporal Lobe Resections in Children with Epilepsy: Neuropsychiatric Status in Relation to Neuropathology and Seizure Outcome. Epilepsy Behav 3, 76–81 (2002). 10.1006/ebeh.2001.0297

79 Datta, A. et al. Selective amygdalohippocampectomy: surgical outcome in children versus adults. Can J Neurol Sci 36, 187–191 (2009).

80 de la Vaissiere, S. et al. Cortical involvement in focal epilepsies with epileptic spasms. Epilepsy Res 108, 1572–1580 (2014). 10.1016/j.eplepsyres.2014.08.008

81 Di Vito, L. et al. Epileptic networks in patients with bitemporal epilepsy: the role of SEEG for the selection of good surgical candidates. Epilepsy Res 128, 73–82 (2016). 10.1016/j.eplepsyres.2016.10.015

82 Diamond, J. M. et al. Travelling waves reveal a dynamic seizure source in human focal epilepsy. Brain 144, 1751–1763 (2021). 10.1093/brain/awab089

83 Ding, P. et al. Contralateral Hippocampal Stimulation for Failed Unilateral Anterior Temporal Lobectomy in Patients with Bilateral Temporal Lobe Epilepsy. Stereotact Funct Neurosurg 94, 327–335 (2016). 10.1159/000449008

84 Dodick, D. W., Cascino, G. D. & Meyer, F. B. Vascular malformations and intractable epilepsy: outcome after surgical treatment. Mayo Clin Proc 69, 741–745 (1994). 10.1016/s0025-6196(12)61091-x

85 Durica, S. R. et al. Stereo-EEG Evaluation and Surgical Treatment in Patients With Drug-Resistant Focal Epilepsy Associated With Nodular Heterotopia. J Clin Neurophysiol 40, 17–26 (2023). 10.1097/WNP.0000000000000850

86 Dylgjeri, S. et al. Insular and insulo-opercular epilepsy in childhood: an SEEG study. Seizure 23, 300–308 (2014). 10.1016/j.seizure.2014.01.008

87 Elias, G. J. B. et al. Impact of Mesial Temporal Lobe Resection on Brain Structure in Medically Refractory Epilepsy. World Neurosurg 152, e652–e665 (2021). 10.1016/j.wneu.2021.06.039

88 Elliott, C. A., Gross, D. W., Wheatley, B. M., Beaulieu, C. & Sankar, T. Progressive contralateral hippocampal atrophy following surgery for medically refractory temporal lobe epilepsy. Epilepsy Res 125, 62–71 (2016). 10.1016/j.eplepsyres.2016.06.007

89 Esquenazi, Y. et al. Surgical Resection for Epilepsy Following Cerebral Gunshot Wounds. World Neurosurg 95, 276–284 (2016). 10.1016/j.wneu.2016.08.041

90 Estupinan-Diaz, B. O. et al. Corpora amylacea in the neocortex in patients with temporal lobe epilepsy and focal cortical dysplasia. Neurologia 30, 90–96 (2015). 10.1016/j.nrl.2013.06.008

91 Fan, X. et al. Optimized SEEG-guided radiofrequency thermocoagulation for mesial temporal lobe epilepsy with hippocampal sclerosis. Seizure 71, 304–311 (2019). 10.1016/j.seizure.2019.08.011

92 Fauser, S. et al. Multi-focal occurrence of cortical dysplasia in epilepsy patients. Brain 132, 2079–2090 (2009). 10.1093/brain/awp145

93 Fierain, A. et al. Stereoelectroencephalography (SEEG) and epilepsy surgery in posttraumatic epilepsy: A multicenter retrospective study. Epilepsy Behav 112, 107378 (2020). 10.1016/j.yebeh.2020.107378

94 Flaus, A. et al. Deep-learning predicted PET can be subtracted from the true clinical fluorodeoxyglucose PET co-registered to MRI to identify the epileptogenic zone in focal epilepsy. Epilepsia Open 8, 1440–1451 (2023). 10.1002/epi4.12820

95 Flaus, A. et al. (18)F-FDG PET/MR in focal epilepsy: A new step for improving the detection of epileptogenic lesions. Epilepsy Res 178, 106819 (2021). 10.1016/j.eplepsyres.2021.106819

96 Fountas, K. N. et al. Temporal pole proton preoperative magnetic resonance spectroscopy in patients undergoing surgery for mesial temporal sclerosis. Neurosurg Focus 32, E3 (2012). 10.3171/2012.1.FOCUS11327

97 Francione, S. et al. Surgery for drug resistant partial epilepsy in children with focal cortical dysplasia: anatomical-clinical correlations and neurophysiological data in 10 patients. J Neurol Neurosurg Psychiatry 74, 1493–1501 (2003). 10.1136/jnnp.74.11.1493

98 Freri, E. et al. Perisylvian, including insular, childhood epilepsy: Presurgical workup and surgical outcome. Epilepsia 58, 1360–1369 (2017). 10.1111/epi.13816

99 Fujiwara, H. et al. Ictal MEG onset source localization compared to intracranial EEG and outcome: improved epilepsy presurgical evaluation in pediatrics. Epilepsy Res 99, 214–224 (2012). 10.1016/j.eplepsyres.2011.11.007

100 Galicia, E. et al. Changing ictal-onset EEG patterns in children with cortical dysplasia. Brain Dev 31, 569–576 (2009). 10.1016/j.braindev.2008.08.011

101 Garganis, K. et al. Frontal lobe epilepsy and mild malformation with oligodendroglial hyperplasia: Further observations on electroclinical and imaging phenotypes, and surgical perspectives. Epileptic Disord 25, 343–359 (2023). 10.1002/epd2.20062

102 Ghods, A. J., Ruban, D. S., Wallace, D. & Byrne, R. W. Differentiating ictal panic with low-grade temporal lobe tumors from psychogenic panic attacks. J Clin Psychiatry 74, 1071–1075 (2013). 10.4088/JCP.13m08378

103 Gnatkovsky, V. et al. Biomarkers of epileptogenic zone defined by quantified stereo-EEG analysis. Epilepsia 55, 296–305 (2014). 10.1111/epi.12507

104 Goldstein, H. E. et al. Safety and efficacy of stereoelectroencephalography in pediatric focal epilepsy: a single-center experience. J Neurosurg Pediatr 22, 444–452 (2018). 10.3171/2018.5.PEDS1856

105 Gonzalez Otarula, K. A. et al. Psychogenic nonepileptic seizures in patients with surgically treated temporal lobe epilepsy: Presurgical and de novo postsurgical occurrence. Epilepsy Behav 75, 252–255 (2017). 10.1016/j.yebeh.2017.08.015

106 Gonzalez-Martinez, J. et al. Stereoelectroencephalography in children and adolescents with difficult-to-localize refractory focal epilepsy. Neurosurgery 75, 258-268; discussion 267-258 (2014). 10.1227/NEU.0000000000000453

107 Gonzalez-Martinez, J. A. et al. The endoscopic anterior transmaxillary temporal pole approach for mesial temporal lobe epilepsies: a feasibility study. J Neurosurg 138, 992–1001 (2023). 10.3171/2022.7.JNS221062

108 Groppel, G. et al. Single stage epilepsy surgery in children and adolescents with focal cortical dysplasia type II - Prognostic value of the intraoperative electrocorticogram. Clin Neurophysiol 130, 20–24 (2019). 10.1016/j.clinph.2018.09.023

109 Gross, R. E. et al. Stereotactic laser amygdalohippocampotomy for mesial temporal lobe epilepsy. Ann Neurol 83, 575–587 (2018). 10.1002/ana.25180

110 Grouiller, F. et al. All-in-one interictal presurgical imaging in patients with epilepsy: single-session EEG/PET/(f)MRI. Eur J Nucl Med Mol Imaging 42, 1133–1143 (2015). 10.1007/s00259-015-3045-2

111 Guan, Y., Zhou, J., Luan, G. & Liu, X. Surgical treatment of patients with Rasmussen encephalitis. Stereotact Funct Neurosurg 92, 86–93 (2014). 10.1159/000355901

112 Guedj, E. et al. 18FDG-PET in different subtypes of temporal lobe epilepsy: SEEG validation and predictive value. Epilepsia 56, 414–421 (2015). 10.1111/epi.12917

113 Gutierrez, C. et al. Clinical features and postoperative seizure outcome in patients with drug-resistant gelastic seizures without hypothalamic hamartoma. Epilepsy Behav 64, 90–93 (2016). 10.1016/j.yebeh.2016.09.038

114 Haegelen, C. et al. High-frequency oscillations, extent of surgical resection, and surgical outcome in drug-resistant focal epilepsy. Epilepsia 54, 848–857 (2013). 10.1111/epi.12075

115 Harroud, A. et al. Precuneal epilepsy: Clinical features and surgical outcome. Epilepsy Behav 73, 77–82 (2017). 10.1016/j.yebeh.2017.05.018

116 Hashiguchi, K. et al. Utility of 3-T FLAIR and 3D short tau inversion recovery MR imaging in the preoperative diagnosis of hippocampal sclerosis: direct comparison with 1.5-T FLAIR MR imaging. Epilepsia 51, 1820–1828 (2010). 10.1111/j.1528-1167.2010.02685.x

117 Heers, M. et al. Monofocal MEG in lesional TLE: does video EEG monitoring add crucial information? Epilepsy Res 92, 54–62 (2010). 10.1016/j.eplepsyres.2010.08.007

118 Ho, S. S., Berkovic, S. F., McKay, W. J., Kalnins, R. M. & Bladin, P. F. Temporal lobe epilepsy subtypes: differential patterns of cerebral perfusion on ictal SPECT. Epilepsia 37, 788–795 (1996). 10.1111/j.1528-1157.1996.tb00653.x

119 Hori, T. et al. Selective subtemporal amygdalohippocampectomy for refractory temporal lobe epilepsy: operative and neuropsychological outcomes. J Neurosurg 106, 134–141 (2007). 10.3171/jns.2007.106.1.134

120 Hudgins, R. J. et al. Surgical treatment of epilepsy in children caused by focal cortical dysplasia. Pediatr Neurosurg 41, 70–76 (2005). 10.1159/000085159

121 Iannotti, G. R. et al. Modulation of epileptic networks by transient interictal epileptic activity: A dynamic approach to simultaneous EEG-fMRI. Neuroimage Clin 28, 102467 (2020). 10.1016/j.nicl.2020.102467

122 Jabbari, B., Prokhorenko, O., Khajavi, K. & Mena, H. Intractable epilepsy and mild brain injury: incidence, pathology and surgical outcome. Brain Inj 16, 463–467 (2002). 10.1080/02699050110102086

123 Jayalakshmi, S., Panigrahi, M., Reddy, R. & Somayajula, S. Clinical characteristics and long-term outcome of surgery for hypothalamic hamartoma in children with refractory epilepsy. Ann Indian Acad Neurol 17, 43–47 (2014). 10.4103/0972-2327.128547

124 Jayapaul, P., Gopinath, S. & Pillai, A. Outcome following surgery for insulo-opercular epilepsies. J Neurosurg 137, 1226–1236 (2022). 10.3171/2021.12.JNS212220

125 Jeong, A. et al. Hemispherotomy in children with electrical status epilepticus of sleep. J Neurosurg Pediatr 19, 56–62 (2017). 10.3171/2016.8.PEDS16319

126 Job, A. S. et al. Epileptogenicity Maps of Intracerebral Fast Activities (60-100 Hz) at Seizure Onset in Epilepsy Surgery Candidates. Front Neurol 10, 1263 (2019). 10.3389/fneur.2019.01263

127 Jobst, B. C. et al. Intractable seizures of frontal lobe origin: clinical characteristics, localizing signs, and results of surgery. Epilepsia 41, 1139–1152 (2000). 10.1111/j.1528-1157.2000.tb00319.x

128 Jobst, B. C. et al. Intractable occipital lobe epilepsy: clinical characteristics and surgical treatment. Epilepsia 51, 2334–2337 (2010). 10.1111/j.1528-1167.2010.02673.x

129 Kaiboriboon, K., Nagarajan, S., Mantle, M. & Kirsch, H. E. Interictal MEG/MSI in intractable mesial temporal lobe epilepsy: spike yield and characterization. Clin Neurophysiol 121, 325–331 (2010). 10.1016/j.clinph.2009.12.001

130 Kalina, M. et al. Stereotactic amygdalohippocampectomy for temporal lobe epilepsy: promising results in 16 patients. Epileptic Disord 9 Suppl 1, S68–74 (2007). 10.1684/epd.2008.0158

131 Kamida, T. et al. The relationship between paired pulse magnetic MEP and surgical prognosis in patients with intractable epilepsy. Seizure 16, 113–119 (2007). 10.1016/j.seizure.2006.10.009

132 Kang, J. Y. et al. Laser interstitial thermal therapy for medically intractable mesial temporal lobe epilepsy. Epilepsia 57, 325–334 (2016). 10.1111/epi.13284

133 Karenfort, M., Kruse, B., Freitag, H., Pannek, H. & Tuxhorn, I. Epilepsy surgery outcome in children with focal epilepsy due to tuberous sclerosis complex. Neuropediatrics 33, 255–261 (2002). 10.1055/s-2002-36740

134 Kazemi, N. J. et al. Resection of frontal encephalomalacias for intractable epilepsy: outcome and prognostic factors. Epilepsia 38, 670–677 (1997). 10.1111/j.1528-1157.1997.tb01236.x

135 Kim, D. W. et al. Parietal lobe epilepsy: the semiology, yield of diagnostic workup, and surgical outcome. Epilepsia 45, 641–649 (2004). 10.1111/j.0013-9580.2004.33703.x

136 Kim, H. I. et al. Bilateral perisylvian ulegyria: clinicopathological study of patients presenting with pseudobulbar palsy and epilepsy. Neuropathology 26, 236–242 (2006). 10.1111/j.1440-1789.2006.00659.x

137 Kim, S. K., Wang, K. C. & Cho, B. K. Intractable seizures associated with brain tumor in childhood: lesionectomy and seizure outcome. Childs Nerv Syst 11, 634–638 (1995). 10.1007/BF00300720

138 Piazzini, A. et al. Visuoperceptive impairment in adult patients with occipital lobe epilepsies. Epilepsy Behav 15, 256–259 (2009). 10.1016/j.yebeh.2009.02.040

139 Rehman, A. & Al Khalili, Y. in StatPearls (2025).

140 Salanova, V., Andermann, F., Rasmussen, T., Olivier, A. & Quesney, L. F. Parietal lobe epilepsy. Clinical manifestations and outcome in 82 patients treated surgically between 1929 and 1988. Brain 118 (Pt 3), 607–627 (1995). 10.1093/brain/118.3.607

141 Thomas, J. et al. Investigating current clinical opinions in stereoelectroencephalography-informed epilepsy surgery. Epilepsia 65, 2662–2672 (2024). 10.1111/epi.18076

142 Barba, C. et al. Seizure Outcome of Temporal Lobe Epilepsy Surgery in Adults and Children: A Systematic Review and Meta-Analysis. Neurosurgery 91, 676–683 (2022). 10.1227/neu.0000000000002094

143 Englot, D. J., Breshears, J. D., Sun, P. P., Chang, E. F. & Auguste, K. I. Seizure outcomes after resective surgery for extra-temporal lobe epilepsy in pediatric patients. J Neurosurg Pediatr 12, 126–133 (2013). 10.3171/2013.5.PEDS1336

144 Krucoff, M. O. et al. Rates and predictors of success and failure in repeat epilepsy surgery: A meta-analysis and systematic review. Epilepsia 58, 2133–2142 (2017). 10.1111/epi.13920

145 Tellez-Zenteno, J. F., Dhar, R. & Wiebe, S. Long-term seizure outcomes following epilepsy surgery: a systematic review and meta-analysis. Brain 128, 1188–1198 (2005). 10.1093/brain/awh449

146 Bjellvi, J., Olsson, I., Malmgren, K. & Wilbe Ramsay, K. Epilepsy duration and seizure outcome in epilepsy surgery: A systematic review and meta-analysis. Neurology 93, e159–e166 (2019). 10.1212/WNL.0000000000007753

147 Tudur Smith, C. et al. Individual participant data meta-analyses compared with meta-analyses based on aggregate data. Cochrane Database Syst Rev 9, MR000007 (2016). 10.1002/14651858.MR000007.pub3

148 Silfvenius, H., Gloor, P. & Rasmussen, T. Evaluation of Insular Ablation in Surgical Treatment of Temporal Lobe Epilepsy. Epilepsia 5, 307–320 (1964). 10.1111/j.1528-1157.1964.tb03338.x

149 Harroud, A., Bouthillier, A., Weil, A. G. & Nguyen, D. K. Temporal lobe epilepsy surgery failures: a review. Epilepsy Res Treat 2012, 201651 (2012). 10.1155/2012/201651

150 Li, J. et al. Insular Involvement in Cases of Epilepsy Surgery Failure. Brain Sci 12 (2022). 10.3390/brainsci12020125

151 Solanki, C., Williams, J., Andrews, C., Fayed, I. & Wu, C. Insula in epilepsy - “untying the gordian knot”: A systematic review. Seizure 106, 148–161 (2023). 10.1016/j.seizure.2023.02.019

152 Obaid, S. et al. Predictors of outcomes after surgery for medically intractable insular epilepsy: A systematic review and individual participant data meta-analysis. Epilepsia Open 8, 12–31 (2023). 10.1002/epi4.12663

153 Harward, S. C., Chen, W. C., Rolston, J. D., Haglund, M. M. & Englot, D. J. Seizure Outcomes in Occipital Lobe and Posterior Quadrant Epilepsy Surgery: A Systematic Review and Meta-Analysis. Neurosurgery 82, 350–358 (2018). 10.1093/neuros/nyx158

154 Jehi, L. et al. Timing of referral to evaluate for epilepsy surgery: Expert Consensus Recommendations from the Surgical Therapies Commission of the International League Against Epilepsy. Epilepsia 63, 2491–2506 (2022). 10.1111/epi.17350

