## Supplementary for "Anatomical Determinants of Epilepsy Surgery Outcomes: A Systematic Review and Individual Patient Data Meta-Analysis"

**Supplementary Materials**

Appendix 1. Detailed search strategy

Date: 8.9.2024

Database (including vendor/platform): MEDLINE (via Ovid)

| Set # | Search Strategy | Results |
| --- | --- | --- |
| 1 Epilepsy | exp Epilepsies, Partial/ OR ((epilepsy.tw. OR epilepsies.tw. OR epileptic.tw. OR epileptics.tw.) adj5 (partial.tw. OR focal.tw. OR occipital.tw. OR digestive.tw. OR abdominal.tw. OR occipital.tw. OR gelastic.tw. OR lobe.tw. OR frontal.tw. OR temporal.tw.)) | 37530 |
| 2 Drug resistant | (Intractable or "medically refractory" or "medication refractory” or "pharmacoresistant" or "pharma-coresistant" or "pharmaco-resistant" or pharmatcoresistance or "pharma-coresistance" or pharmaco-resistance or ((treatment or treatments or therapy or therapies or therapeutic or therapeutics or therapeutically or drug or drugs or medication or medications or medically) adj3 resist*)). tw. | 244974 |
| 3 Surgery | surgery.fs. OR exp "Neurosurgical Procedures"/ OR neurosurg*.tw. OR lobectomy.tw. OR lobectomies.tw. OR surgery.tw. OR surgeries.tw. OR surgical.tw. OR procedure.tw. OR procedures.tw. OR procedural.tw. OR operation.tw. OR operations.tw. OR operative.tw. OR resection.tw. OR resections.tw. OR resectioned.tw. OR resectioning.tw. OR thermocoagulation.tw. OR ablation.tw. OR ablations.tw. | 4677765 |
| 4 Treatment Outcome | exp "Treatment Outcome"/ OR outcome.tw. OR outcomes.tw. OR (Engel.tw. adj3 (score.tw. OR scores.tw. OR scored.tw. OR scoring.tw.)) | 3139318 |
| 5 | 1 and 2 and 3 and 4 | 2074 |
|  | 11484687 OR 29069568 OR 33452051 OR 29768433 | 1/4 |

Database: Embase (Elsevier)

| Set # | Search Strategy | Results |
| --- | --- | --- |
| 1 Epilepsy | 'focal epilepsy'/exp OR ((epilepsy OR epilepsies OR epileptic OR epileptics) NEAR/5 (partial OR focal OR occipital OR digestive OR abdominal OR occipital OR gelastic OR lobe OR frontal OR temporal)):ti,ab | 73,578 |
| 2 Drug resistant | Intractable:ti,ab OR 'medically refractory':ti,ab OR 'medication refractory':ti,ab OR pharmacoresistant:ti,ab OR pharma-coresistant:ti,ab OR pharmaco-resistant:ti,ab OR pharmatcoresistance:ti,ab OR pharma-coresistance:ti,ab OR pharmaco-resistance:ti,ab OR ((treatment OR treatments OR therapy OR therapies OR therapeutic OR therapeutics OR therapeutically OR drug OR drugs OR medication OR medications OR medically) NEAR/3 resist*):ti,ab | 349,521 |
| 3 Surgery | 'Surgery'/de OR 'Neurosurgery'/exp OR neurosurg*:ti,ab OR lobectomy:ti,ab OR lobectomies:ti,ab OR surgery:ti,ab OR surgeries:ti,ab OR surgical:ti,ab OR procedure:ti,ab OR procedures:ti,ab OR procedural:ti,ab OR operation:ti,ab OR operations:ti,ab OR operative:ti,ab OR resection:ti,ab OR resections:ti,ab OR resectioned:ti,ab OR resectioning:ti,ab OR thermocoagulation:ti,ab OR ablation:ti,ab OR ablations:ti,ab | 5,573,328 |
| 4 Treatment Outcome | 'Treatment Outcome'/exp OR outcome:ti,ab OR outcomes:ti,ab OR (Engel NEAR/3 (score OR scores OR scored OR scoring)):ti,ab | 4,533,210 |
| 5 | #1 AND #2 AND #3 AND #4 | 3,427 |
| 6 | #1 AND #2 AND #3 AND #4 AND [humans]/lim | 3237 |

Database: Scopus (Elsevier)

| Set # | Search Strategy | Results |
| --- | --- | --- |
| 1 Epilepsy | TITLE-ABS({focal epilepsy} OR ((epilepsy OR epilepsies OR epileptic OR epileptics) W/5 (partial OR focal OR occipital OR digestive OR abdominal OR occipital OR gelastic OR lobe OR frontal OR temporal))) | 33,669 |
| 2 Drug resistant | TITLE-ABS(Intractable OR {medically refractory} OR {medication refractory} OR pharmacoresistant OR pharma-coresistant OR pharmaco-resistant OR pharmatcoresistance OR pharma-coresistance OR pharmaco-resistance OR ((treatment OR treatments OR therapy OR therapies OR therapeutic OR therapeutics OR therapeutically OR drug OR drugs OR medication OR medications OR medically) W/3 resist*)) | 340,342 |
| 3 Surgery | TITLE-ABS(neurosurg* OR lobectomy OR lobectomies OR (surgery OR surgeries OR surgical OR procedure OR procedures OR procedural OR operation OR operations OR operative) OR resection OR resections OR resectioned OR resectioning OR thermocoagulation OR ablation OR ablations) | 7,760,969 |
| 4 Treatment Outcome | TITLE-ABS(outcome OR outcomes OR (Engel W/3 (score OR scores OR scored OR scoring))) | 3,107,212 |
| 5 | #1 AND #2 AND #3 AND #4 | 1,705 |

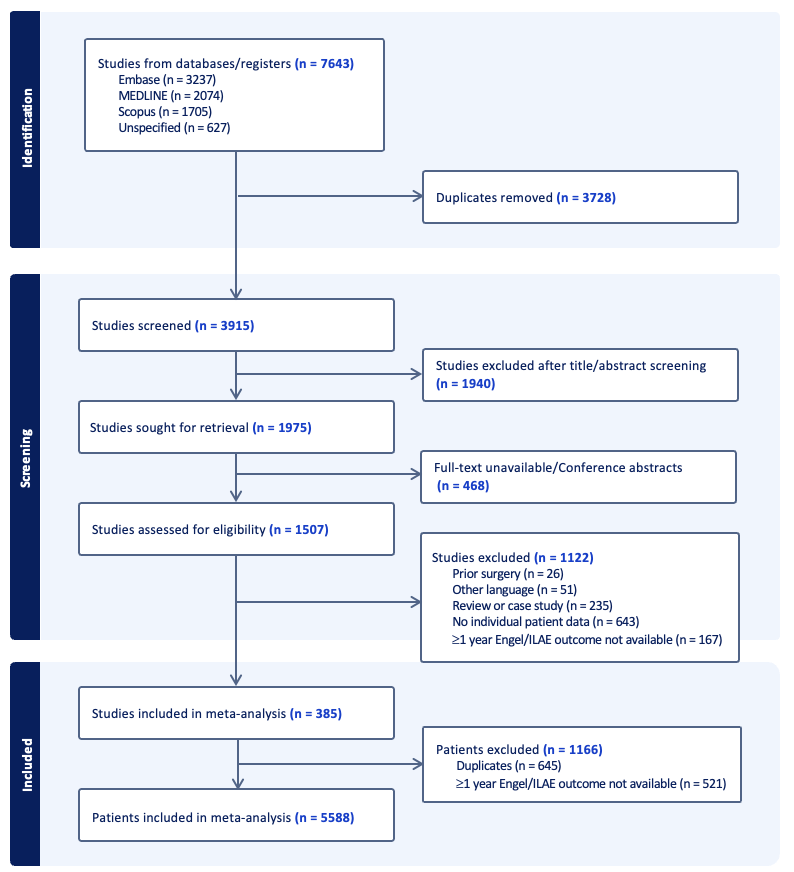

Supplementary Figure 1. PRISMA flow diagram for individual participant data. To identify relevant patient cohorts that aligned with the scope of this meta-analysis, the following inclusion criteria were applied: (i) original research involving human subjects, (ii) availability of individual patient-level information, (iii) availability of at least 1-year post-surgical outcomes (1-year is the minimum accepted follow-up time frame in epilepsy surgery outcome assessment), and (iv) specification of epileptic focus or type of epilepsy. We excluded reviews and case studies (n<3), as well as articles that were not written in English, French, Spanish, or Mandarin.

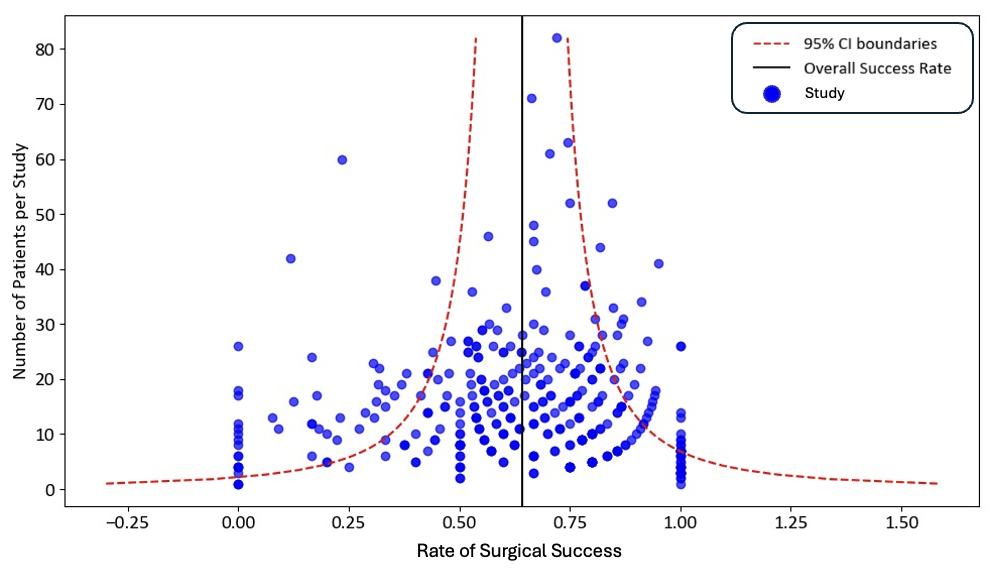

Supplementary Figure 2. Funnel plot to assess publication bias. Visual observation of the plot reveals a slight asymmetry.

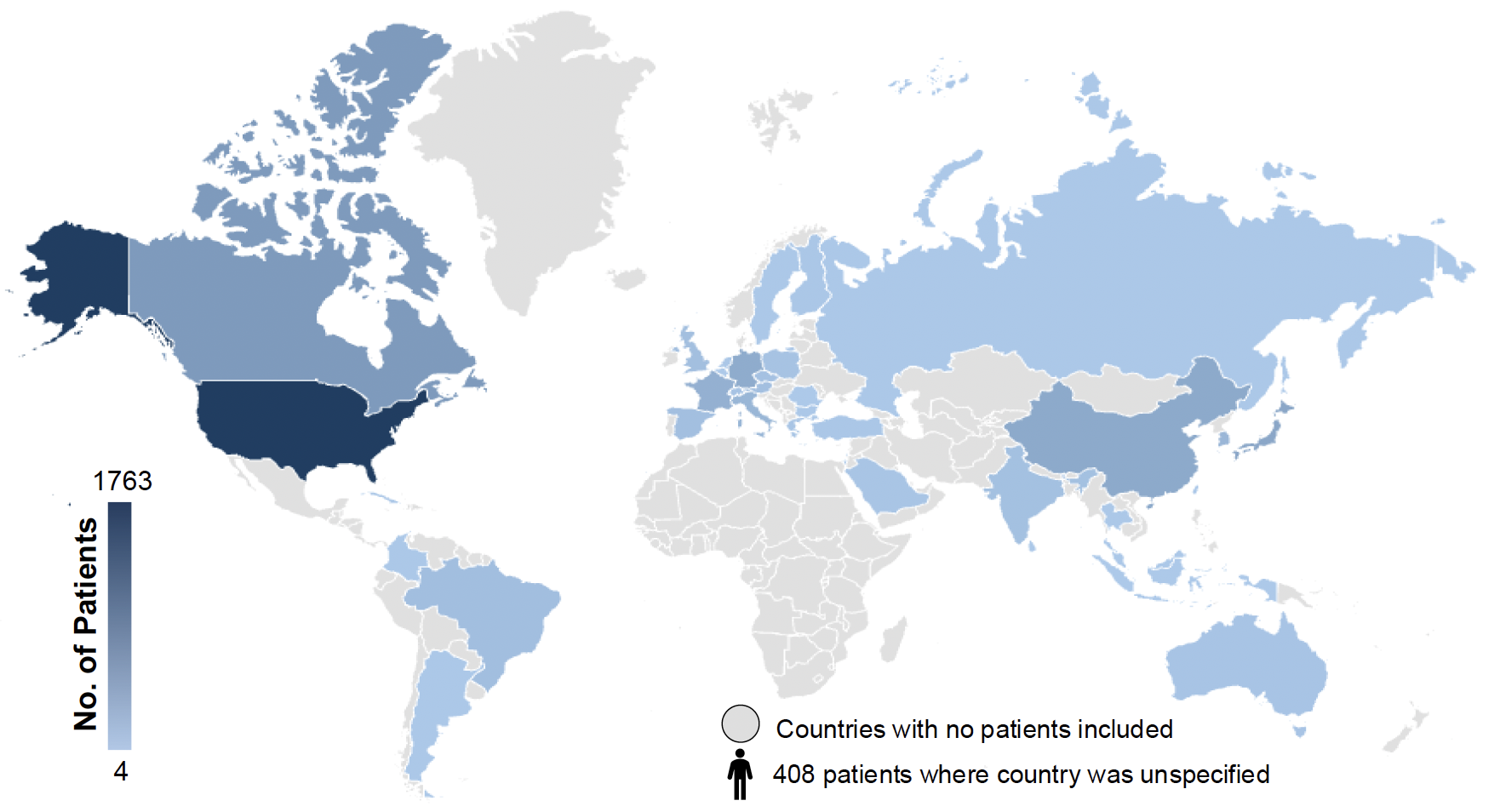

Supplementary Figure 3. Heatmap representing the geographical distribution of included patients. Our meta-analysis encompassed patients from 36 countries, listed in descending order based on the number of patients included: United States of America, Canada, Japan, China, Germany, France, Italy, South Korea, Brazil, United Kingdom, Austria, Spain, India, Czech Republic, Poland, Switzerland, Australia, Greece, Saudi Arabia, Netherlands, Cuba, Finland, Turkey, Israel, Colombia, Argentina, Taiwan, Thailand, Bulgaria, Russia, Sweden, Malaysia, Sri Lanka, Belgium, Romania, Indonesia.

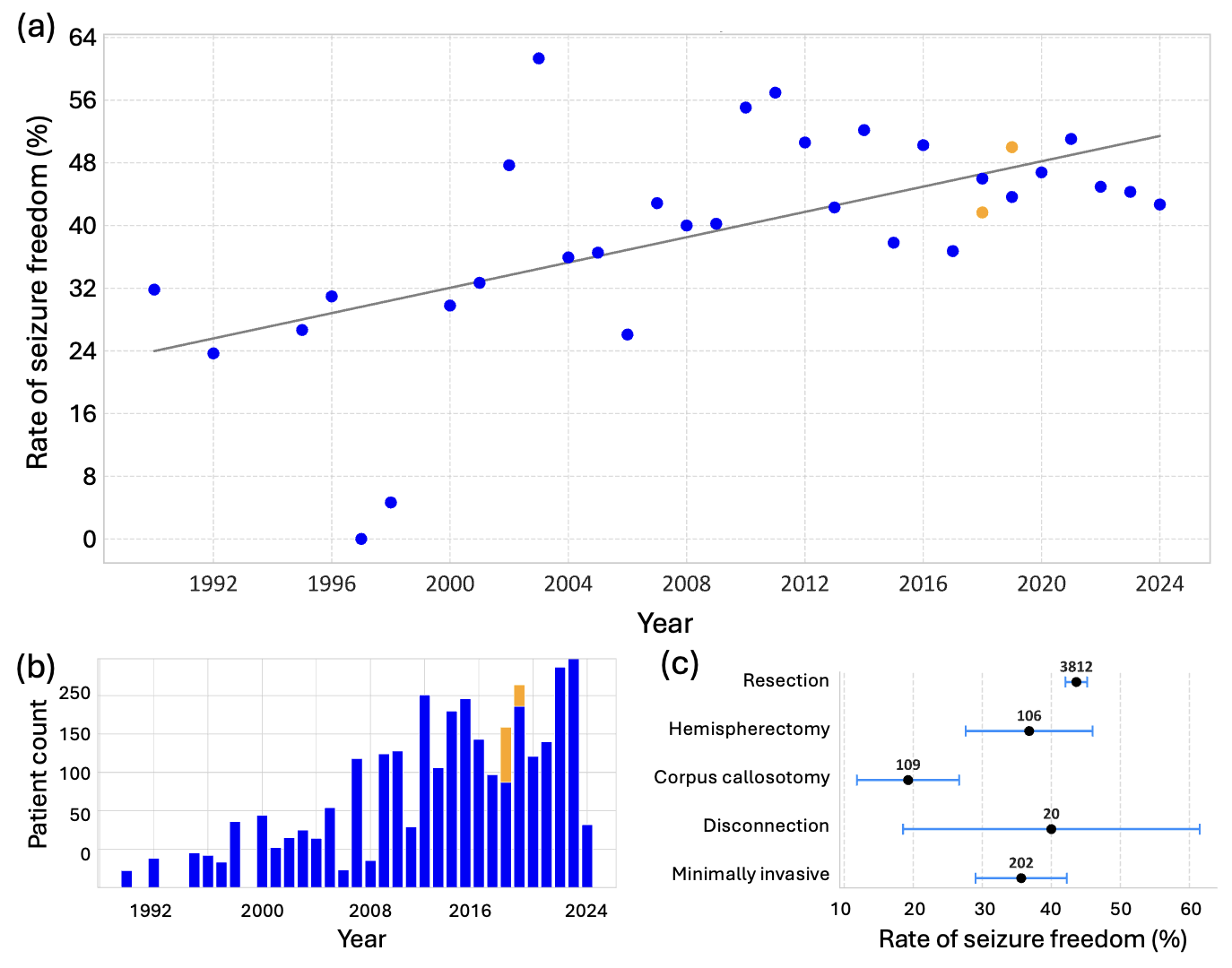

Supplementary Figure 4. Type of surgical interventions. (a) Reported rates of seizure freedom (blue) (defined as Engel 1a/ILAE 1) over time, along with the rates of seizure freedom for minimally invasive surgeries (orange). Each dot represents the overall rate of seizure freedom calculated from all the patients pooled across studies conducted in a given year. Linear regression revealed a significant trend (r=0.59, p<0.01). (b) Number of patients undergoing resections and those receiving minimally invasive surgeries over time. Cases of Engel 1 where the subgroup were not specified were excluded. (c) Seizure freedom rates by intervention type were as follows: resection (43.6% [42.1-45.2%]), hemispherectomy (36.8% [27.6-46.0%]), corpus callosotomy (19.3% [11.9-26.7%]), disconnection (40.0% [18.5-61.5%]), and minimally invasive surgery (35.6% [29.0-42.2%]). Significant differences were observed between the following pairs: hemispherectomy vs. corpus callosotomy (p<0.01, V=0.18), corpus callosotomy vs. resection (p<0.01, V=0.08), corpus callosotomy vs. minimally invasive surgery (p<0.01, V=0.16), and resection vs. minimally invasive surgery (p=0.03, V=0.03).

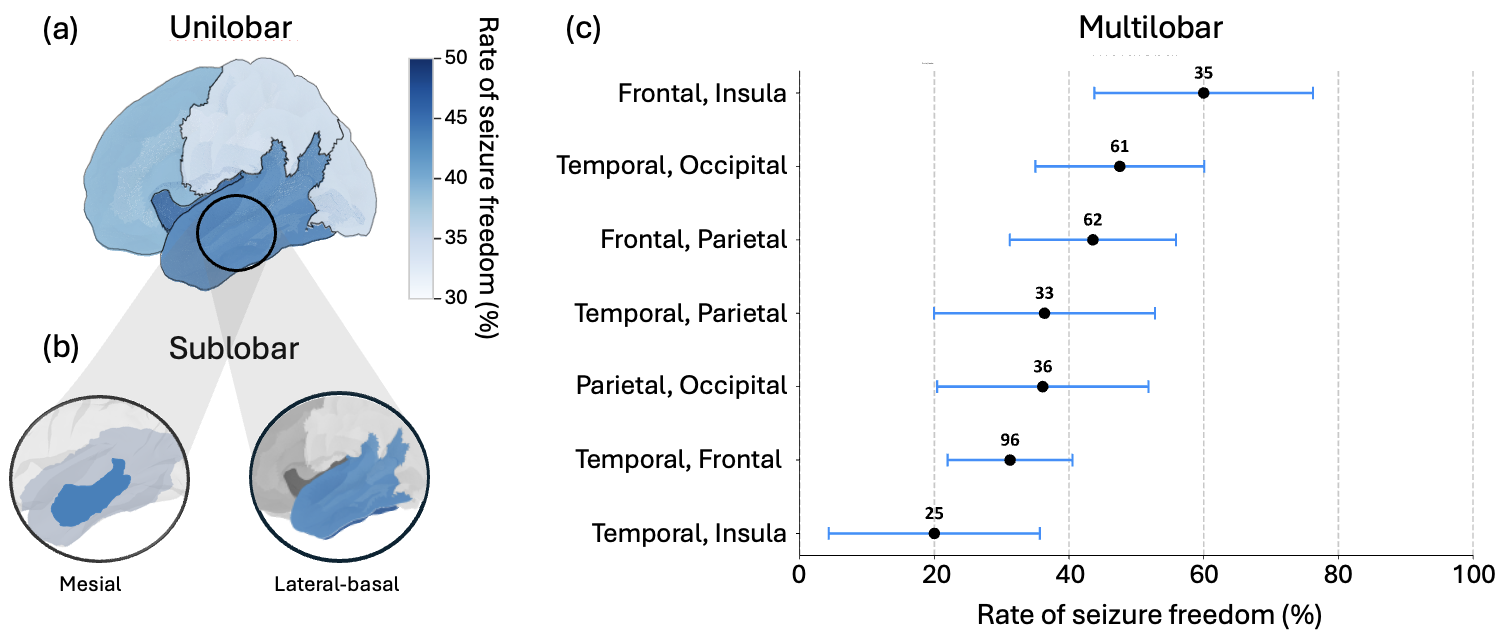

Supplementary Figure 5. Rates of seizure freedom (Engel 1a/ILAE 1) across unilobar, sublobar and multilobar surgeries. (a) For unilobar surgeries, seizure freedom rates varied significantly by lobe (χ²=14.6, p<0.01, V=0.06). The highest success rates were observed in the insula (n=58; 48.3% [35.4–61.1%]), followed by the temporal lobe (n=2480; 46.0% [44.0–47.9%]) and frontal lobe (n=589; 40.2% [36.3–44.2%]). The lowest success rates were seen in the parietal (n=157; 35.7% [28.2–43.2%]) and occipital lobes (n=90; 35.6% [25.7–45.4%]). In paired chi-square tests, only temporal vs. frontal and temporal vs. parietal comparisons reached statistical significance. (b) Within the temporal lobe, sublobar analysis showed that mesial temporal surgeries (n=469) had a success rate of 43.9% (39.4–48.4%), whereas procedures targeting the lateral and basal temporal regions (n=70) had a success rate of 51.4% (39.7–63.1%). (c) For multilobar surgeries, there was a significant association between surgical region and success rate (χ²=25, p=0.02, V=0.22). Success rates for different lobe combinations were as follows: frontal-insula (60.0% [43.8–76.2%]), temporal-occipital (47.5% [35.0–60.1%]), frontal-parietal (43.5% [31.2–55.9%]), temporal-parietal (36.4% [20.0–52.8%]), parietal-occipital (36.1% [20.4–51.8%]), temporal-frontal (31.3% [22.0–40.5%]), and temporal-insula (20.0% [4.3–35.7%]). In paired chi-square tests, significant differences were observed between frontal-insula vs. temporal-frontal, temporal-occipital vs. temporal-insula, and temporal-insula vs. frontal-insula.

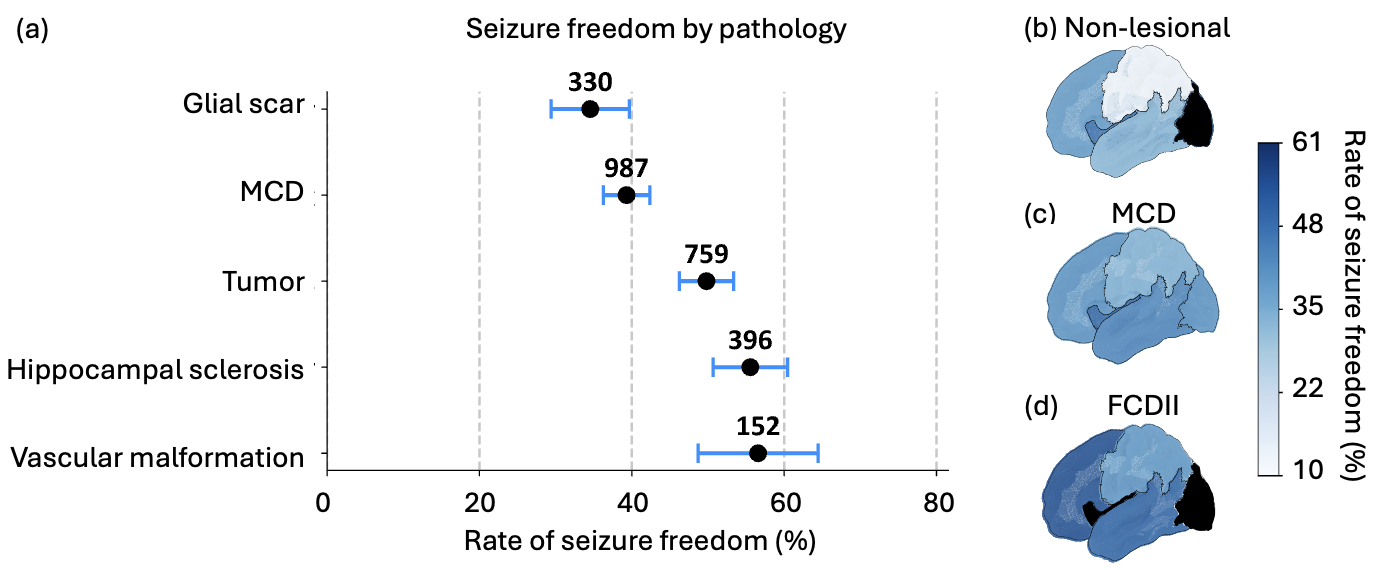

Supplementary Figure 6. Rates of seizure freedom (Engel Ia/ILAE 1) by pathology. (a) Rates of seizure freedom were significantly associated with pathology (χ²=60.5, p<0.01, V=0.15). The success rates for the top five pathologies in our cohort were as follows: vascular malformation (56.6% [48.7–64.5%]), tumors (55.6% [50.7–60.4%]), hippocampal sclerosis (49.8% [46.2–53.4%]), malformation of cortical development (39.3% [36.3–42.4%]), and glial scars (34.5% [29.4–39.7%]). There was no significant association between seizure freedom rates and the lobe involved in (b) MRI negative epilepsy (n=403, χ²=8.5, p=0.09, V=0.14), (c) malformation of cortical development (n=807, χ²=3.6, p=0.45, V=0.06), or (d) focal cortical dysplasia (FCD) type II (n=315, χ²=3.8, p=0.49, V=0.08).

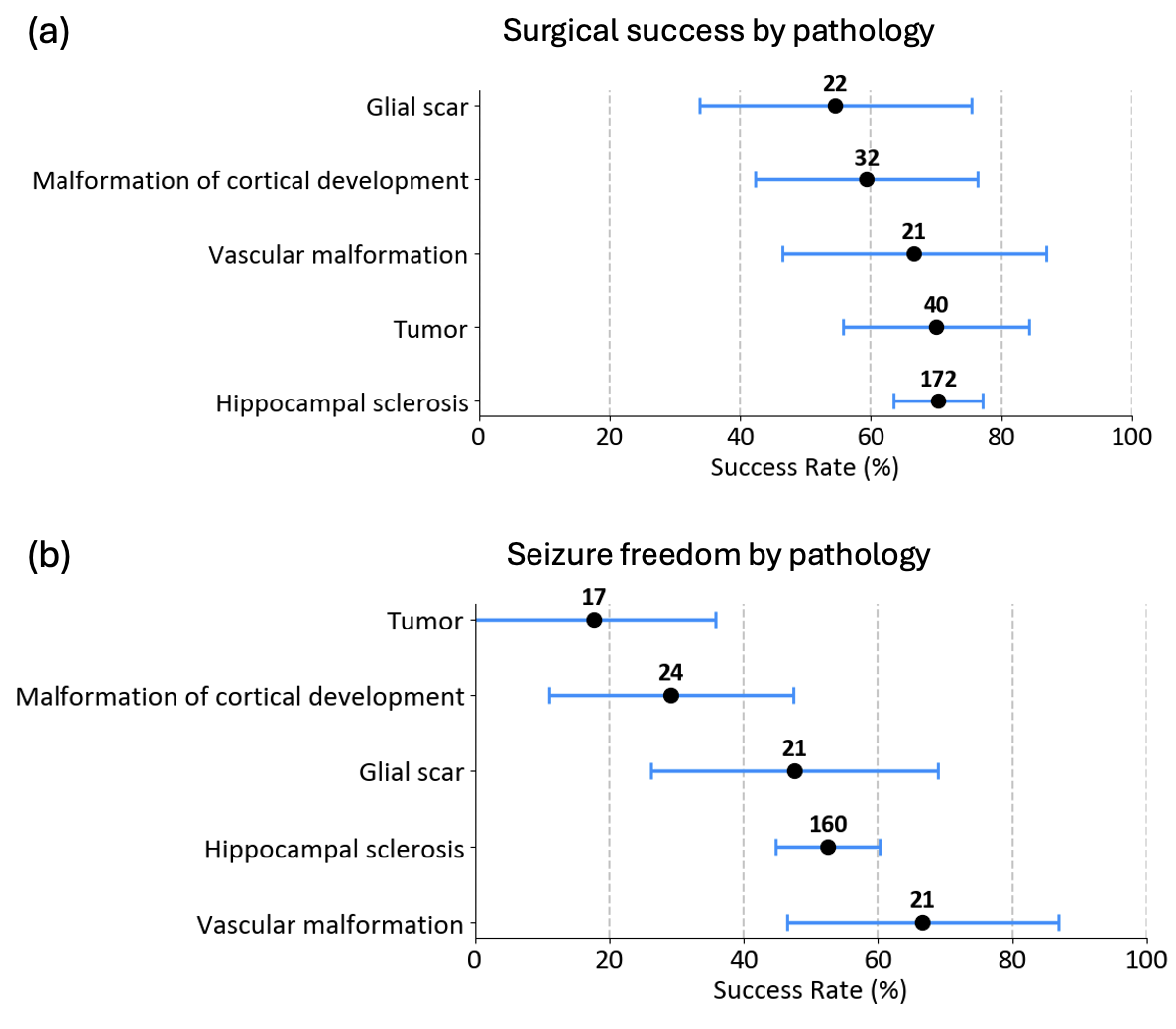

Supplementary Figure 7. Rates of (a) surgical success and (b) seizure freedom by pathology in mesial temporal lobe epilepsy.

Supplementary Table 1. Description of included studies.

| **Source** | **Country** | **Sample Size** | **Age Group** | **Scale Used for Outcomes** | **Prevalence of Good Outcomes** | **Prevalence of Seizure Freedom** |
| --- | --- | --- | --- | --- | --- | --- |
| Abdallah et al., 2022 | Canada | 15 | Adult | Engel classification | 40% | 33% |
| Abel et al., 2018 | France | 12 | Pediatric | Engel classification | 33% | 33% |
| Aghakani et al., 2005 | Canada | 6 | Adult | Engel classification | 67% | 33% |
| Akeret et al., 2018 | Switzerland | 15 | Adult + Pediatric | Engel classification | 73% | 67% |
| Alsaadi et al., 2003 | United States | 15 | Adult | Engel classification | 87% | 60% |
| Alshafi et al., 2014 | Canada | 4 | Pediatric | Engel classification | 25% | 0% |
| Arévalo-Astrada et al., 2021 | Canada | 6 | Adult | Engel classification | 67% | 0% |
| Aung et al., 2021 | United States | 4 | Pediatric | Engel classification | 100% | 100% |
| Aznarez et al., 2020 | Spain | 19 | Adult | Engel classification | 68% | 58% |
| Bakhtiar et al., 2022 | Indonesia | 4 | Adult | Engel classification | 75% | 0% |
| Bansal et al., 2016 | United States | 14 | Pediatric | Engel classification | 50% | 0% |
| Barba et al., 2013 | Multiple | 12 | Adult + Pediatric | Engel classification | 67% | 67% |
| Bauer et al., 2007 | Austria | 14 | Adult | Engel classification | 86% | 75% |
| Benedetti-Isaac et al., 2013 | Colombia | 21 | Adult + Pediatric | Engel classification | 76% | 43% |
| Benifla et al., 2017 | Israel | 22 | Pediatric | Engel classification | 82% | 0% |
| Benifla et al., 2009 | Canada | 22 | Pediatric | Engel classification | 64% | 0% |
| Blauwblomme et al., 2013 | France | 16 | Adult + Pediatric | Engel classification | 81% | 63% |
| Boling, 2010 | United States | 17 | Adult | Engel classification | 71% | 0% |
| Boling et al., 2009 | Canada | 11 | Adult | Engel classification | 55% | 0% |
| Bourgeois et al., 2007 | France | 26 | Pediatric | Engel classification | 81% | 69% |
| Buchfelder et al., 2002 | Germany | 27 | Adult + Pediatric | Engel classification | 74% | 67% |
| Buraniqi et al., 2023 | United States | 10 | Pediatric | Engel classification | 70% | 50% |
| Burneo et al., 2003 | United States | 5 | Adult | Engel classification | 100% | 0% |
| Busch et al., 2008 | United States | 12 | Adult | Engel classification | 83% | 67% |
| Caicoya et al., 2007 | Spain | 7 | Adult | Engel classification | 71% | 57% |
| Cajigas et al., 2019 | United States | 29 | Adult | Engel classification | 62% | 41% |
| Cao et al., 2022 | China | 31 | Adult | Engel classification | 77% | 77% |
| Carreño et al., 2018 | Multiple | 13 | Adult + Pediatric | Engel classification | 15% | 0% |
| Casadei et al., 2020 | United States | 13 | Adult + Pediatric | Engel classification | 8% | 0% |
| Casciatio et al., 2015 | Italy | 5 | Adult | Engel classification | 40% | 20% |
| Catarino et al., 2011 | Multiple | 9 | Adult | ILAE classification | 89% | 89% |
| Centeno et al., 2017 | United Kingdom | 20 | Pediatric | ILAE classification | 55% | 50% |
| Chandra et al., 2008 | India | 19 | Pediatric | Engel classification | 95% | 0% |
| Chang et al., 2009 | United States | 13 | Adult + Pediatric | Engel classification | 38% | 0% |
| Chassoux, 2003 | France | 3 | Adult | Engel classification | 100% | 100% |
| Chen et al., 2014 | Taiwan | 8 | Pediatric | Engel classification | 75% | 0% |
| Chen et al., 2015 | United States | 7 | Pediatric | ILAE classification | 100% | 86% |
| Chen et al., 2005 | Germany | 22 | Adult | Engel classification | 86% | 68% |
| Choi et al., 2013 | South Korea | 5 | Adult | Engel classification | 80% | 0% |
| Choi et al., 2003 | South Korea | 47 | Adult + Pediatric | Engel classification | 81% | 49% |
| Chugani et al., 2014 | United States | 23 | Pediatric | ILAE classification | 83% | 74% |
| Cimbalnik et al., 2019 | Multiple | 50 | Adult | ILAE classification and Engel Classification | 52% | 48% |
| Citherlet et al., 2021 | Canada | 15 | Adult | Engel classification | 67% | 0% |
| Cohen-Gadol et al., 2003 | United States | 5 | Adult | Engel classification | 80% | 0% |
| Cossu et al., 2005 | Italy | 33 | Pediatric | Engel classification | 58% | 39% |
| Cossu et al., 2014 | Italy | 4 | Adult + Pediatric | Engel classification | 100% | 75% |
| Costa et al., 2022 | Belgium | 7 | Adult | Engel classification | 86% | 0% |
| D'Agostino et al., 2004 | Multiple | 12 | Adult + Pediatric | Engel classification | 42% | 0% |
| D'Alessio et al., 2014 | Argentina | 14 | Adult | Engel classification | 50% | 0% |
| Daniel et al., 2007 | Multiple | 13 | Adult + Pediatric | Engel classification | 92% | 85% |
| Danielsson et al., 2002 | Sweden | 13 | Pediatric | Engel classification | 54% | 0% |
| Datta et al., 2009 | Canada | 23 | Pediatric | Engel classification | 57% | 0% |
| de la Vaissière et al., 2014 | France | 10 | Pediatric | Engel classification | 60% | 40% |
| Di Gennaro et al., 2015 | Italy | 6 | Adult | Engel classification | 17% | 0% |
| Di Vito et al., 2016 | France | 7 | Adult | Engel classification | 86% | 71% |
| Diamond et al., 2021 | United States | 20 | Adult | Engel classification | 70% | 50% |
| Ding et al., 2016 | China | 18 | Adult | Engel classification | 33% | 0% |
| Dodick et al., 1994 | United States | 20 | Adult | Engel classification | 75% | 0% |
| Durica et al., 2023 | United States | 6 | Adult | Engel classification | 67% | 67% |
| Dylgjeri et al., 2014 | France | 9 | Pediatric | Engel classification | 78% | 0% |
| Elias et al., 2021 | Canada | 19 | Adult | Engel classification | 74% | 0% |
| Elliott et al., 2016 | Canada | 26 | Adult | Engel classification | 77% | 73% |
| Esquenazi et al., 2016 | United States | 4 | Adult | Engel classification | 50% | 25% |
| Estupiñán-Díaz et al., 2015 | Cuba | 8 | Adult | Engel classification | 63% | 63% |
| Fan et al., 2019 | China | 21 | Adult | Engel classification | 76% | 38% |
| Fauser et al., 2009 | Germany | 4 | Adult | Engel classification | 100% | 100% |
| Fierain et al., 2020 | France | 12 | Adult | Engel classification | 67% | 0% |
| Flaus et al., 2023 | France | 13 | Adult | Engel classification | 54% | 46% |
| Flaus et al., 2021 | France | 17 | Adult | Engel classification | 76% | 65% |
| Fountas et al., 2012 | Greece | 23 | Adult | Engel classification | 74% | 0% |
| Freri et al., 2017 | Italy | 16 | Pediatric | Engel classification | 56% | 44% |
| Fujiwara et al., 2011 | United States | 6 | Pediatric | Engel classification | 50% | 0% |
| Galicia et al., 2009 | Canada | 12 | Pediatric | Engel classification | 67% | 50% |
| Garganis et al., 2023 | Greece | 5 | Adult + Pediatric | Engel classification | 60% | 40% |
| Ghods et al., 2013 | United States | 10 | Adult | Engel classification | 80% | 70% |
| Gil et al., 2010 | Argentina | 6 | Adult + Pediatric | Engel classification | 67% | 30% |
| Gnatkovsky et al., 2014 | Italy | 12 | Adult + Pediatric | Engel classification | 83% | 83% |
| Goldstein et al., 2018 | United States | 12 | Pediatric | Engel classification | 50% | 0% |
| González Otárula et al., 2017 | Multiple | 16 | Adult | Engel classification | 56% | 44% |
| Gonzalez-Martinez et al., 2014 | United States | 18 | Pediatric | Engel classification | 56% | 0% |
| Gonzalez-Martinez et al., 2022 | United States | 4 | Adult | Engel classification | 100% | 100% |
| Gröppel et al., 2019 | Austria | 18 | Pediatric | ILAE classification | 72% | 72% |
| Gross et al., 2018 | United States | 49 | Adult | Engel classification | 57% | 46% |
| Grouiller et al., 2015 | Switzerland | 4 | Adult + Pediatric | Engel classification | 75% | 0% |
| Guan et al., 2014 | China | 20 | Pediatric | Engel classification | 80% | 0% |
| Guedj et al., 2015 | France | 30 | Adult + Pediatric | Engel classification | 53% | 7% |
| Gutierrez et al., 2016 | United States | 9 | Adult | Engel classification | 67% | 0% |
| Haegelen et al., 2013 | Canada | 22 | Adult | ILAE classification | 32% | 18% |
| Hajek et al., 1991 | Switzerland | 25 | Adult + Pediatric | Engel classification | 52% | 0% |
| Harroud et al., 2017 | Canada | 6 | Adult | Engel classification | 33% | 33% |
| Hashiguchi et al., 2010 | Japan | 13 | Adult | Engel classification | 77% | 0% |
| Heers et al., 2010 | Germany | 25 | Adult | Engel classification | 80% | 48% |
| Hirabayashi et al., 1992 | United Kingdom | 17 | Adult + Pediatric | Engel classification | 18% | 0% |
| Ho et al., 1996 | Australia | 30 | Adult | Engel classification | 80% | 53% |
| Hori et al., 2007 | Japan | 26 | Adult | Engel classification | 54% | 20% |
| Hudgins et al., 2005 | United States | 15 | Pediatric | Engel classification | 67% | 67% |
| Hyslop et al., 2015 | United States | 25 | Pediatric | Engel classification | 60% | 0% |
| Iannotti et al., 2020 | Multiple | 3 | Pediatric | Engel classification | 33% | 0% |
| Jabbari et al., 2009 | United States | 9 | Adult | Engel classification | 89% | 89% |
| Jayalakshmi et al., 2014 | India | 6 | Pediatric | Engel classification | 50% | 0% |
| Jayapaul et al., 2022 | India | 22 | Adult + Pediatric | Engel classification | 73% | 55% |
| Jeong et al., 2017 | United States | 8 | Pediatric | Engel classification | 100% | NA |
| Job et al., 2019 | France | 17 | Adult + Pediatric | Engel classification | 35% | 29% |
| Jobst et al., 2000 | United States | 25 | Adult + Pediatric | Engel classification | 64% | 48% |
| Jobst et al., 2010 | United States | 12 | Adult | Engel classification | 50% | 0% |
| Juhász et al., 2010 | United States | 20 | Pediatric | Engel classification | 70% | 0% |
| Juhász et al., 2001 | United States | 15 | Adult + Pediatric | Engel classification | 53% | 53% |
| Jung et al., 2013 | Austria | 16 | Adult | ILAE classification | 0% | 0% |
| Kaiboriboon et al., 2010 | United States | 22 | Adult | Engel classification | 77% | 68% |
| Kalina et al., 2007 | Czech Republic | 16 | Adult | Engel classification | 75% | 0% |
| Kamida et al., 2007 | Japan | 5 | Adult + Pediatric | Engel classification | 80% | 0% |
| Kang et al., 2015 | United States | 12 | Adult | Engel classification | 75% | 0% |
| Karenfort et al., 2002 | Germany | 4 | Pediatric | Engel classification | 50% | 50% |
| Kashida et al., 2019 | Japan | 18 | Adult | Engel classification | 94% | 67% |
| Kazemi et al., 1997 | United States | 17 | Adult + Pediatric | Engel classification | 59% | 0% |
| Kim et al., 2013 | United States | 14 | Pediatric | Engel classification | 86% | 0% |
| Kim et al., 2006 | Korea | 5 | Adult | Engel classification | 20% | 0% |
| Kim et al., 1995 | Korea | 10 | Pediatric | Engel classification | 60% | 0% |
